# Climate-Driven Malaria Transmission Dynamics with Human Awareness and Optimal Control: A Deterministic Mathematical Modeling Approach

**DOI:** 10.64898/2026.07.29.26359260

**Authors:** Robert Nyamao Nyabwanga, Lewis Ketter, Abraham N. Osogo, Robert Karieko Obogi, Lameck Ondieki Agasa, Fred Nyamitago Monari

## Abstract

Malaria is still one of the most dangerous causes of morbidity and mortality in tropical and subtropical regions even though it has been actively combated for many years. In 2023, there were approximately 263 million malaria cases and 597,000 deaths from this disease on a global scale, with sub-Saharan Africa being the region most affected by it [24]. Climate factors affect mosquito biology, including their abundance, survival, and biting rates, as well as parasite development, while human awareness plays a crucial role in adopting preventive measures and effective treatments. Despite the progress in both climate- and awareness-based malaria modelings, few studies integrate these factors in one comprehensive model that involves the detailed mechanisms of transmission processes. The current study develops a deterministic climate-driven SEAIR-SEI malaria transmission model that includes the impact of temperature, rainfall, and humidity on mosquito biology and endogenous community awareness. The model was proven to be well-posed by showing the positivity and boundedness of its solution and through the demonstration of the existence and uniqueness of its solution. The malaria-free equilibrium was determined, and the basic reproduction number was calculated using the next-generation matrix method. The model underwent local and global stability analyses to characterise the disease’s persistence in the population. Additionally, a normalized forward sensitivity analysis was conducted, revealing the mosquito biting rate as the key force driving malaria transmission. Four time-dependent malaria interventions, namely, long-lasting insecticidal nets, community awareness campaigns, indoor residual spraying, and prompt treatment, were included in the model through optimal control theory and analysed using Pontryagin’s Maximum Principle. The numerical results for the optimal control problem showed that employing all four interventions leads to the best outcome by decreasing the objective functional value by 88.17%, reducing the total number of infected humans by 92.49%, and minimizing the total number of infectious mosquitoes by 93.87%. Interestingly, combining two interventions, indoor residual spraying, and prompt treatment, also yielded nearly optimal results. Therefore, the designed control strategy can serve as an efficient and affordable framework for malaria control in sub-Saharan Africa.

## 1 Introduction

Malaria is among the most fatal vector-borne diseases and a major public health problem despite the considerable control and elimination efforts [5, 23, 24]. According to the World Health Organization (WHO), there were 263 million malaria cases and 597,000 malaria deaths worldwide in 2023; 95% of cases and 50% of deaths were reported from the WHO African region [24]. In addition, children under 5 years of age bear the highest risk of malaria mortality, comprising approximately 3 out of every 4 malaria deaths [24]. Nonetheless, significant progress toward malaria control and elimination has been made in recent years due to the extensive use of long-lasting insecticidal nets, indoor residual spraying, malaria rapid diagnostic tests, seasonal malaria chemoprevention, artemisinin-based combination therapy, and improved surveillance [23, 24]. Despite these interventions, sub-Saharan Africa has been experiencing recurrent and persistent transmission of Plasmodium parasites due to the complexity of the association between humans, P. falciparum, the vector, and the environment as well as socio-economic and behavioural drivers [10, 14]. In other words, the links between key factors and the spread of the disease are not static but rather dynamic processes that make it challenging to predict and control malaria transmission [10, 14]. Recently, studies have emphasized the importance of understanding the interactions between social, economic, demographic, environmental, and ecological drivers of malaria transmission at the local level [2, 8, 10].

Unlike most other directly transmitted infections, malaria parasites have an indirect transmission route that requires both human and mosquito hosts to complete the parasite life cycle [20, 23]. As such, malaria epidemiology is inseparable from the ecology of *Anopheles* mosquitoes and the demographic and behavioural characteristics of human populations [14, 23]. Consequently, fluctuations in vector population dynamics, parasite development, climatic conditions, and socioeconomic or demographic variables can substantially influence malaria incidence and prevalence [2, 10, 14]. For example, socio-economic status, maternal education, nutritional status, health-seeking behaviour, the use of insecticide-treated nets, breastfeeding, and residential environmental conditions are important risk factors for malaria infection and severity in young children [8]. Similarly, climate forcing has emerged as a critical driver of malaria transmission since rainfall and temperature strongly influence mosquito abundance, biting rates, survival, and parasite development [1, 17]. Climate-dependent models of malaria transmission have gained considerable traction because they provide mechanistic insights into the interactions among temperature, mosquito ecology, and parasite development [1, 13, 17]. For instance, [13] developed a mechanistic transmission model showing that malaria transmission peaks at approximately 25°C, lower than previously estimated, and declines at higher temperatures because mosquito survival decreases. Similar conclusions were reached by [1], highlighting the importance of incorporating biologically realistic temperature-dependent response functions into malaria transmission models. These findings illustrate the complexity of climate’s effect on malaria transmission and the importance of correctly capturing such mechanisms using realistic temperature-dependent response functions [1, 13, 17].

Rainfall patterns are also critical climate drivers that can impact malaria transmission through their effect on mosquito breeding habitats. For example, rainfall creates aquatic habitats suitable for mosquito larvae, while droughts destroy these habitats, thus decreasing mosquito production [1]. Moreover, rainfall can lead to extreme flood conditions that may drown mosquito larvae, reducing the overall mosquito population. High atmospheric humidity promotes mosquito survival by reducing desiccation rates and extending the lifespan of adult mosquitoes, increasing the likelihood of surviving the parasite’s extrinsic incubation period (EIP). In this context, EIP is the period of time the parasite takes to develop inside mosquitoes after they have fed on a blood meal from an infected human; only mosquitoes that survive this period can transmit the infection. Thus, relatively small changes in humidity, rainfall, and mosquito survival could significantly alter the transmission intensity of the disease and the reproduction number.

It is well established that climate change will have a significant impact on the transmission dynamics of many infectious diseases, including malaria. Changes in temperature and rainfall regimes, atmospheric humidity, and the increased frequency of extreme weather events may affect the geographic distribution and temporal variation in malaria transmission. For instance, recent studies conducted in Ethiopia revealed that rainfall, relative humidity, and temperature were strongly associated with malaria incidence and highlighted the importance of climate-sensitive malaria surveillance systems and early warning systems [1]. On the other hand, several recent compartmental models incorporate seasonality and climate variability in evaluating malaria transmission dynamics and control [6, 13, 15]. Nonetheless, many climate-dependent malaria transmission models only incorporate seasonality through the use of time-dependent periodic transmission coefficients while ignoring the fact that mosquito demographics, biting rates, EIP, and mosquito survival time are affected differently by temperature, rainfall, and humidity.

In addition to climate variability, human behaviour is a significant non-biological driver of malaria transmission. Human behavioural responses to the threat of malaria infection and the availability of prevention and treatment measures are crucial in determining the transmission intensity of the disease [3, 12].

For example, consistent use of treated mosquito nets, environmental management, appropriate treatment-seeking behaviour, and adherence to control measures lower the risk of infection and promote rapid recovery. On the other hand, failure to seek timely treatment and poor treatment adherence have the opposite effect on the disease’s persistence and spread. Awareness campaigns have long been recognized as pivotal to reducing the incidence of various infectious diseases by encouraging individuals to modify their behaviour to avoid infection or seek care when sick [3, 12]. In particular, the work of [3] demonstrated that awareness campaigns, treatment, and the use of insecticides could significantly reduce disease prevalence. They found that awareness campaigns reduced disease prevalence by promoting preventive measures while also increasing treatment rates and insecticide use levels. Furthermore, the study showed that optimal control of these interventions could lower the reproduction number below unity, ensuring effective disease management and elimination.

It is worth noting that awareness is not a static variable; rather, it varies dynamically depending on disease prevalence, personal experience, and awareness campaigns in the local community. As a result, when disease prevalence rises, so does community awareness, leading to increased use of preventive measures such as mosquito nets, environmental management, and treatment-seeking behaviour. Likewise, when prevalence declines, so does awareness, leading to a decrease in preventive measures, creating an association between disease prevalence and awareness levels. This suggests that disease prevalence levels may regulate awareness levels within the local population, contributing to disease persistence and intensifying epidemics. Recently, several researchers have investigated the impact of awareness campaigns and the resulting human behavioural changes on the dynamics of infectious diseases, particularly those transmitted by vectors [3, 15]. Most of these studies demonstrate that awareness campaigns and subsequent changes in individual behavioural patterns have a significant impact on the course of an epidemic.

It is now well understood that climate change and human non-biological factors such as awareness campaigns can significantly influence the dynamics of malaria transmission. Specifically, warmer temperatures and increased humidity promote mosquito proliferation and survival, increasing the density of infective vectors and the risk of infection [1]. Moreover, rising infection rates are likely to increase community awareness and promote the adoption of preventive measures such as using mosquito nets and undergoing treatment, thus reducing the prevalence of infection. On the other hand, effective malaria control measures may lower awareness levels, leading to decreased use of preventive measures such as mosquito nets and treatment-seeking behaviour, leaving populations vulnerable to subsequent climate-driven outbreaks.

Therefore, models that overlook either climate-dependent biological response functions or awareness dynamics and optimal control strategies may fail to capture essential transient dynamics and effective control measures.

Mathematical models have become increasingly useful in understanding the epidemiological dynamics of infectious diseases and developing control strategies. The classic Ross-Macdonald model [11, 18] has undergone numerous extensions and modifications to incorporate the complex interactions between host demographics, mosquito biology, climate, seasonality, immunity, treatment seeking, insecticide use, and optimal control in malaria control [4, 7, 20]. More importantly, sensitivity analysis has been used to assess the relative importance of model parameters in shaping outbreak dynamics [7]. Recent works have expanded on the understanding of mosquito-borne disease dynamics and control by incorporating climate, community awareness, seasonality, insecticide resistance, data assimilation, and optimal control in malaria transmission models [3, 6, 15]. However, most of these studies have failed to explicitly account for both awareness dynamics driven by disease prevalence and mosquito climate-dependent response functions while implementing multiple control interventions.

Motivated by these considerations, the current study develops a non-autonomous climate-driven SEAIR-SEI malaria transmission model that explicitly considers community awareness and optimal control strategies. Specifically, this work makes several novel contributions by incorporating the following key features:

i. We propose a compartmental malaria transmission model with non-autonomous climate-dependent response functions to highlight the importance of realistic temperature, rainfall, and humidity-dependent biological response functions in modulating malaria transmission.
ii. We incorporate awareness dynamics to demonstrate how awareness campaigns influence human behavioural responses, such as mosquito net use, environmental management, and treatment-seeking, to reduce disease prevalence.
iii. We investigate the impact of optimal control strategies, including mosquito net use, awareness campaigns, IRS, and treatment, on curtailing disease transmission.

In this model, the human population is heterogeneous and consists of unaware susceptible, aware susceptible, exposed, infectious, and recovered individuals, with aware susceptible denoting individuals taking preventive measures such as use of mosquito nets, IRS treatment and environmental management. The human population is exposed to malaria infection either through mosquito bites or contaminated human blood transfusion; mosquitoes, on the other hand, acquire the infection through blood meals from infectious humans, with both mosquito and human populations exhibiting the exposed-infectious delay. Temperature, rainfall, and relative humidity are incorporated into the model as explanatory variables via biological response functions that regulate mosquito biting rates, recruitment, survival rates, mosquito-to-human transmission probabilities, and the extrinsic incubation period of the parasite.

## 2 Model Formulation

We formulate a deterministic climate-driven malaria transmission model with dynamic community awareness and integrated control interventions. The human population is partitioned into unaware susceptible 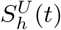, aware susceptible 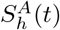, exposed *E*_*h*_(*t*), infectious *I*_*h*_(*t*), and recovered *R*_*h*_(*t*) individuals. The mosquito population comprises susceptible *S*_*m*_(*t*), exposed *E*_*m*_(*t*), and infectious *I*_*m*_(*t*) mosquitoes. Thus,

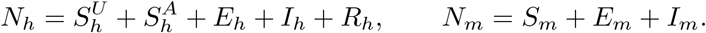

Community malaria awareness is denoted by *A*(*t*), and the complete state vector is

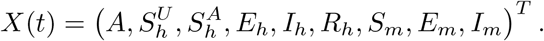

### Model Assumptions

The model assumes that newly recruited humans and mosquitoes enter their respective susceptible classes. Malaria transmission occurs exclusively through mosquito-human interactions, while exposed humans and mosquitoes are infected but not yet infectious. Aware susceptible individuals adopt preventive practices and consequently experience a lower infection risk than unaware individuals.

Community awareness is maintained through routine health education and increases in response to human and mosquito infections and targeted public-health campaigns, but declines without continued reinforcement. Recovered humans acquire temporary immunity and may become susceptible again following immunity loss. Temperature, rainfall, and relative humidity modify malaria transmission and mosquito population dynamics. LLINs reduce mosquito-to-human transmission, IRS increases mosquito mortality, and treatment accelerates human recovery. All state variables and parameters are assumed non-negative.

### Community Awareness Dynamics

Community awareness evolves according to

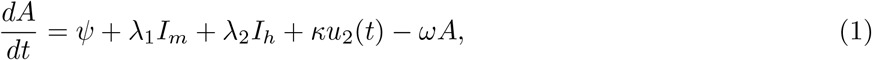

where *ψ* is the baseline awareness input, *λ*_1_ and *λ*_2_ measure infection-driven awareness responses, *κu*_2_(*t*) represents campaign-induced awareness, and *ω* is the awareness decay rate.

The awareness-mediated behavioural transitions are

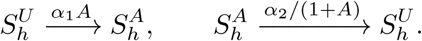

Hence, unaware susceptible individuals acquire protective behaviour at rate 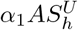, whereas aware individuals lose such behaviour at rate 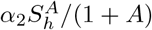. The latter decreases as community awareness increases, representing greater persistence of preventive behaviour.

### Climate-Dependent Biological Response Functions

Let *T* (*t*), *R*(*t*), and *H*(*t*) denote temperature, rainfall, and relative humidity, respectively. Seasonal climate variability is represented by

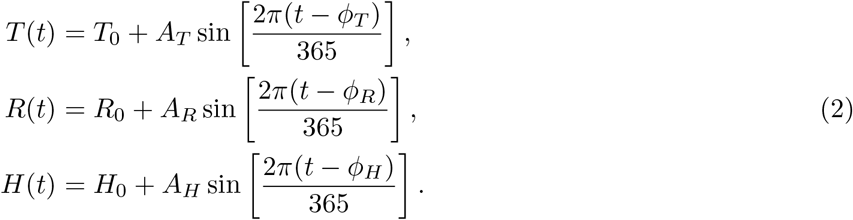

The corresponding normalized climatic deviations are

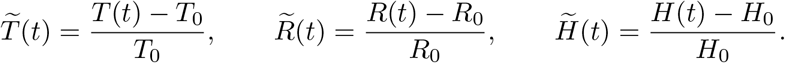

Climate influences mosquito biting activity, mosquito-to-human transmission, mosquito recruitment, mosquito mortality, and the extrinsic incubation rate. These biological responses are modelled as

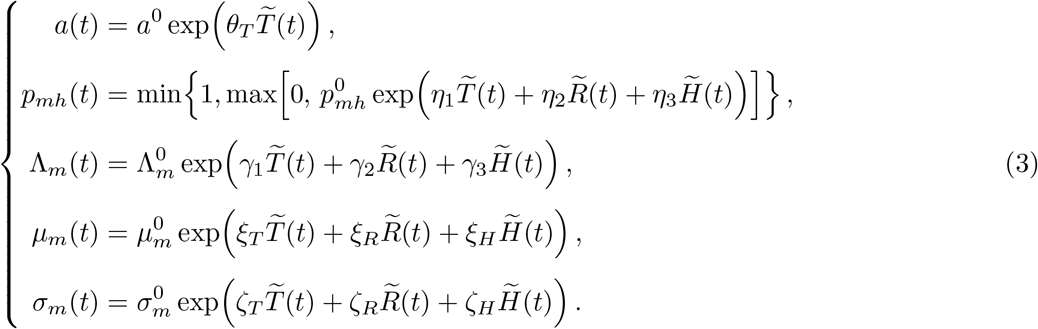

The truncation of *p*_*mh*_(*t*) ensures that

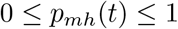

for all *t*. When the climatic variables attain their reference values,

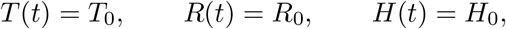

the normalized deviations vanish, yielding

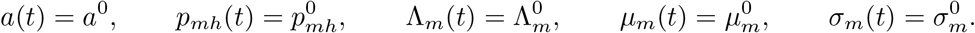

Thus, all climate-dependent biological processes reduce to their respective reference values, and the model recovers the corresponding reference-climate autonomous system.

### Forces of Infection

The mosquito-to-human and human-to-mosquito forces of infection are defined by

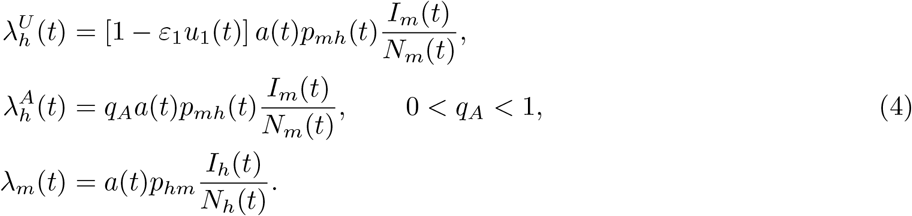

Here, *u*_1_(*t*) denotes LLIN coverage, *ε*_1_ is LLIN effectiveness, *q*_*A*_ represents the reduced relative infection risk among aware susceptible individuals, and *p*_*hm*_ is the human-to-mosquito transmission probability per infectious bite. The climate-dependent biting rate *a*(*t*) therefore couples both directions of vector-host transmission.

### Control Interventions

Four time-dependent malaria interventions are incorporated through the control vector

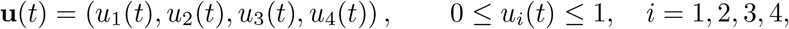

where *u*_1_(*t*) represents LLIN coverage and effective use, *u*_2_(*t*) denotes targeted awareness-campaign intensity, *u*_3_(*t*) represents IRS intensity, and *u*_4_(*t*) denotes treatment effort directed towards infectious humans.

The controls act on distinct transmission and recovery mechanisms. LLINs reduce mosquito-to-human transmission through the factor 1 *− ε*_1_*u*_1_(*t*) in *λ*^*U*^ (*t*), whereas awareness campaigns increase community awareness through *κu*_2_(*t*). IRS increases the effective mosquito mortality rate from *µ*_*m*_(*t*) to *µ*_*m*_(*t*) + *Δu*_3_(*t*), where *Δ* is the maximum IRS-induced mortality rate. Similarly, treatment increases the recovery rate of infectious humans from *ρ*_2_ to *ρ*_2_ + *γu*_4_(*t*), where *γ* denotes the maximum treatment-induced recovery rate.

### Governing Equations

Combining the epidemiological, behavioural, climatic, and intervention mechanisms described above yields the climate-driven SEAIR–SEI malaria transmission model

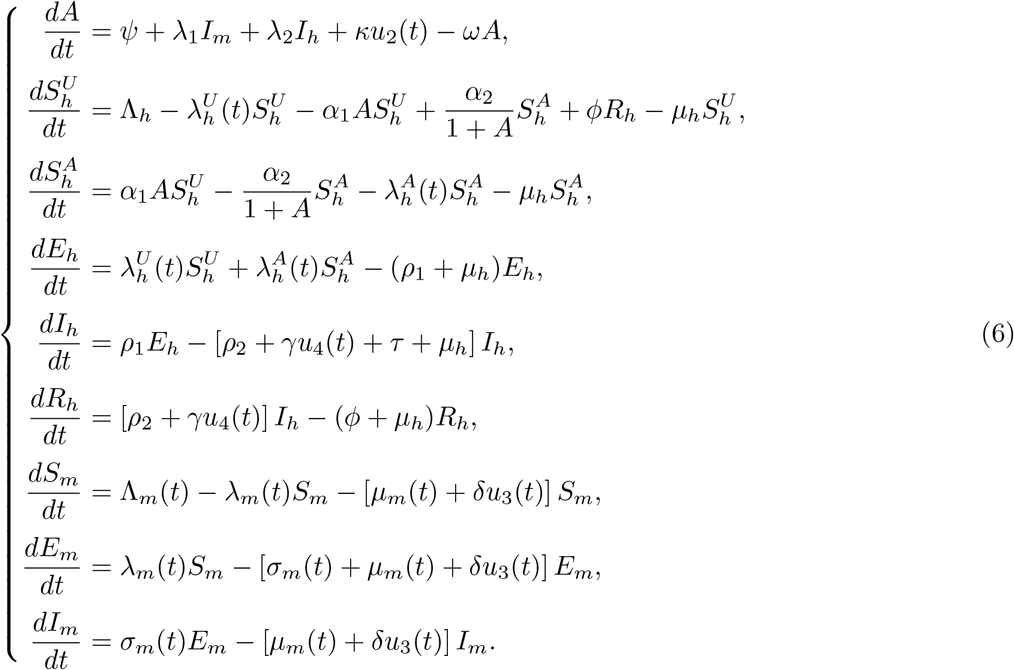

The state variables follow the ordering

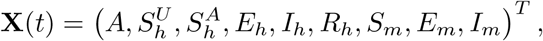

and is adopted in the numerical implementation.

The model is supplemented with the non-negative initial conditions

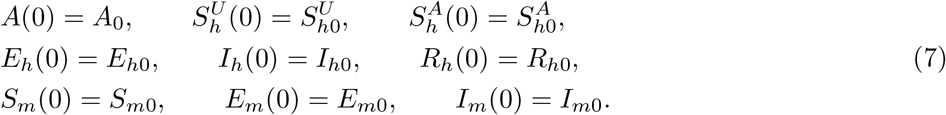

#### AUTONOMOUS Model for Threshold Analysis

The complete system (6) is non-autonomous because seasonal variations in temperature, rainfall, and relative humidity induce time-dependent biological responses. For equilibrium and threshold analyses, the climatic conditions are frozen at their reference values and the controls are set to zero:

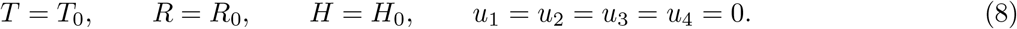

The corresponding biological quantities are denoted by

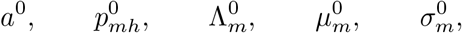

representing the biting rate, mosquito-to-human transmission probability, mosquito recruitment, mosquito mortality, and extrinsic incubation rate at the reference climatic conditions, respectively. The autonomous forces of infection therefore reduce to

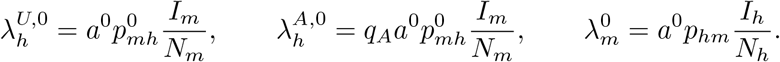

At the disease-free state,

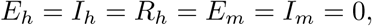

and the awareness equation gives

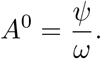

Thus, a positive baseline awareness level permits the coexistence of unaware and aware susceptible humans in the absence of malaria. The resulting autonomous system forms the basis for the disease-free equilibrium, reproduction-number, stability, and bifurcation analyses. The full time-dependent system is retained for seasonal simulations and optimal control analysis. The state variables and parameter descriptions are highlighted in Table 1.

**Table 1.**
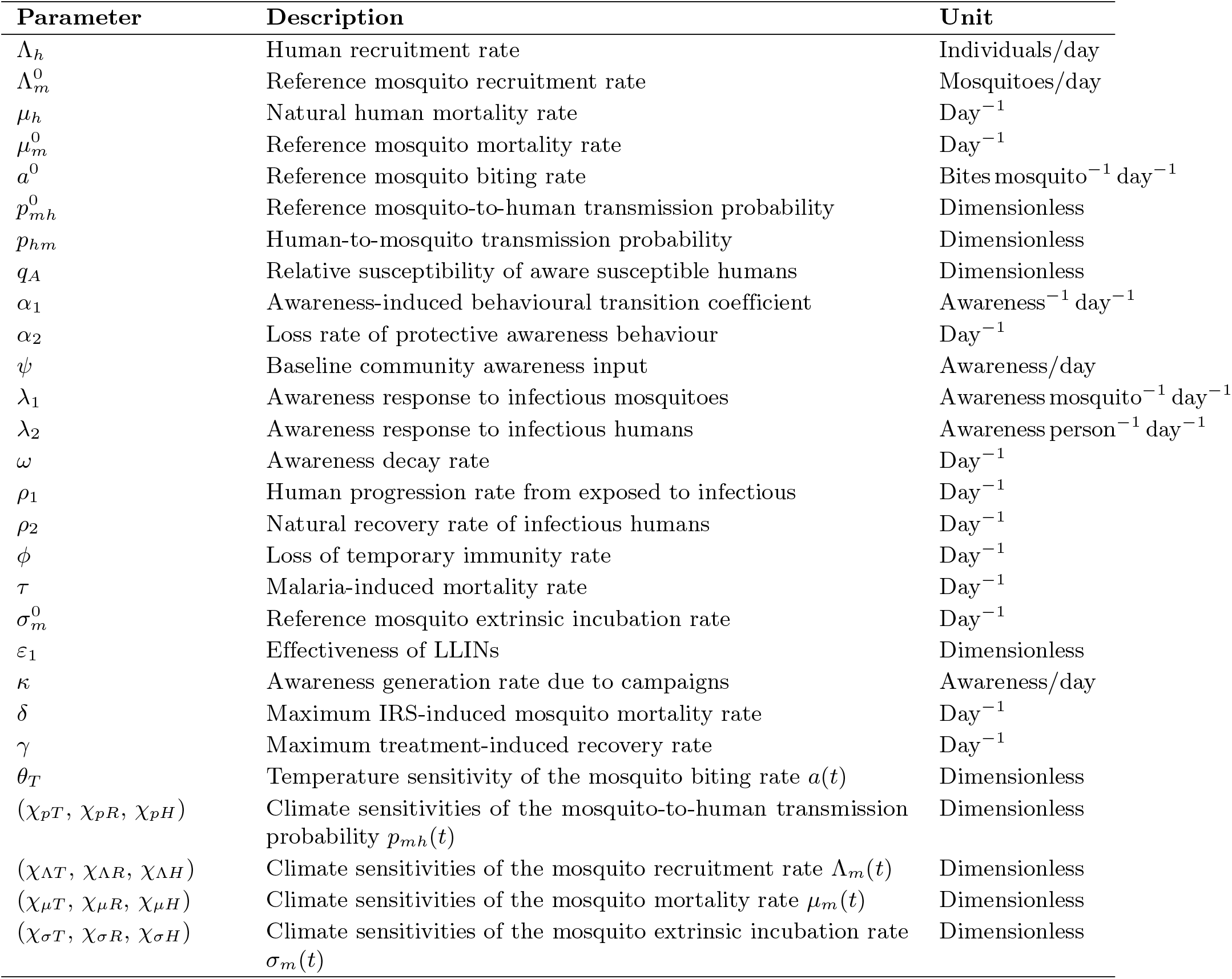
Principal parameters of the climate-driven malaria transmission model.

The epidemiological, behavioural, climatic, and intervention pathways of the model are summarized in Figure 1. The framework integrates human and mosquito infection dynamics, awareness-mediated behavioural transitions, climate-dependent vector processes, and four, time-dependent malaria control interventions.

**Fig 1.**
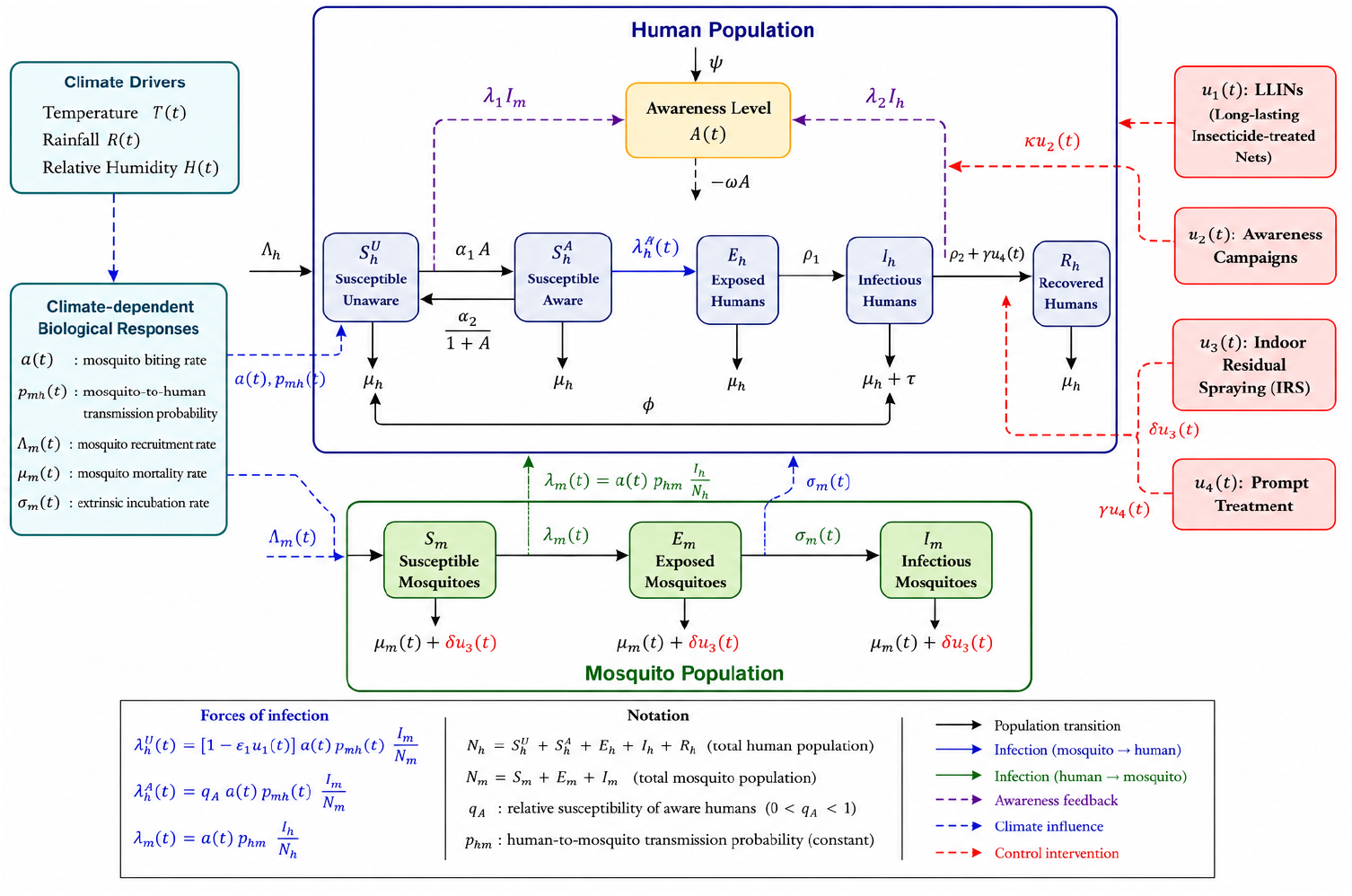
Compartmental structure of the climate-driven SEAIR–SEI malaria transmission model with dynamic community awareness and integrated control interventions. The human population comprises unaware susceptible 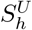, aware susceptible 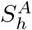, exposed *E*_*h*_, infectious *I*_*h*_, and recovered *R*_*h*_ individuals, while *A*(*t*) represents community awareness. The mosquito population is divided into susceptible *S*_*m*_, exposed *E*_*m*_, and infectious *I*_*m*_ classes. Climatic conditions modify vector biological processes and transmission, while *u*_1_(*t*), *u*_2_(*t*), *u*_3_(*t*), and *u*_4_(*t*) represent LLIN use, awareness campaigns, IRS, and treatment, respectively.

## 3 Mathematical Properties of the Model

### 3.1 Well-Posedness of the Model

We first establish the epidemiological well-posedness of system (6). In particular, solutions originating from non-negative initial conditions must remain non-negative and bounded.

Let

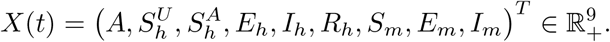

We assume *N*_*h*_(0) *>* 0, *N*_*m*_(0) *>* 0, and bounded continuous climatic functions.

#### Theorem 1

(Positivity of Solutions). *For every non-negative initial condition* 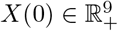, *the solution of system* (6) *remains non-negative for all t ≥* 0.

*Proof*. Evaluating the vector field on the boundary of the non-negative orthant gives

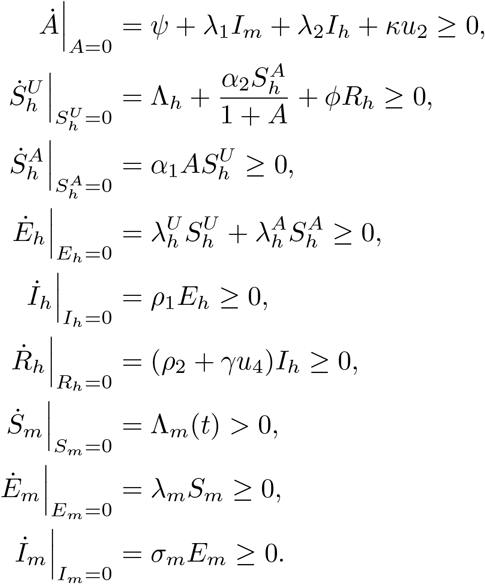

Hence, the vector field is inward-pointing or tangent on each boundary of R^9^ . Therefore, the non-negative orthant is positively invariant.

#### Definition 1

(Boundedness and Feasible Region). *Summing the human equations gives*

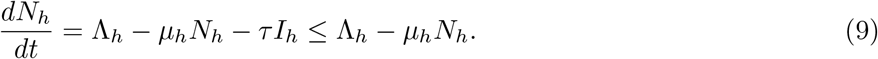

*By the comparison theorem*,

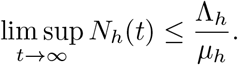

*Similarly, the mosquito population satisfies*

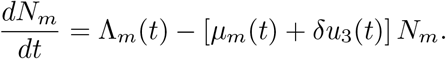

*Since the climatic functions are bounded, define*

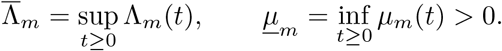

*It follows that*

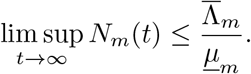

*Moreover, since I*_*h*_ *≤ N*_*h*_, *I*_*m*_ *≤ N*_*m*_, *and* 0 *≤ u*_2_ *≤* 1, *the awareness equation satisfies*

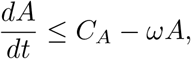

*where*

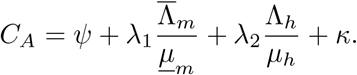

*Consequently*,

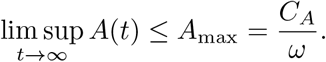

*The epidemiologically feasible region is therefore*

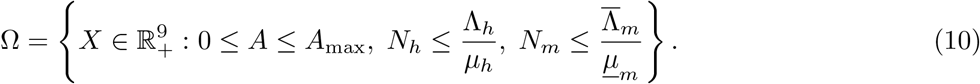

#### Theorem 2

*The region* Ω *is positively invariant for X*(0) ∈ Ω, *and every solution originating from finite non-negative initial data is ultimately bounded*.

*Proof*. Positivity follows from Theorem 1. On the upper boundaries of Ω,

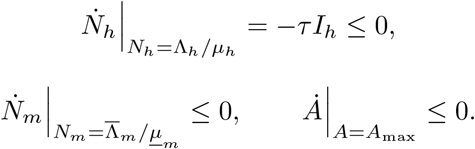

Thus, the vector field cannot point outward through the boundary of Ω. The preceding comparison inequalities establish ultimate boundedness.

#### Theorem 3

(Existence and Uniqueness). *Considering the compact form X*? = *F* (*t, X, u*), *X*(0) = *X*_0_ *and suppose that T* (*t*), *R*(*t*), *and H*(*t*) *are continuous and bounded on every finite time interval and that the controls u*_*i*_(*t*) *are bounded and measurable,then the system* (6) *admits a unique global non-negative solution for every biologically admissible initial condition*.

*Proof*. On the biologically relevant domain *N*_*h*_ *>* 0 and *N*_*m*_ *>* 0, the right-hand side *F* (*t, X, u*) is locally Lipschitz in *X* and satisfies the standard Carathéodory conditions. Hence, a unique local solution exists. Positivity and boundedness prevent finite-time escape or blow-up, so the solution extends globally for all *t ≥* 0.

### 3.2 Malaria-Free Equilibrium and Basic Reproduction Number

The basic reproduction number *R*_0_ represents the expected number of secondary malaria infections generated by a typical infectious individual introduced into an otherwise infection-free population. It is the principal threshold quantity governing malaria invasion and persistence. Unless otherwise stated, the analysis is based on the frozen-climate autonomous system defined in Section 2.

#### Definition 2

(Malaria-Free Equilibrium). *At the malaria-free equilibrium (MFE)*,

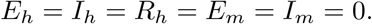

*The awareness equation gives*

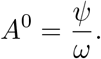

*The susceptible human equilibrium equations are*

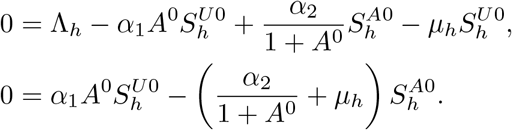

*Define the awareness ratio*

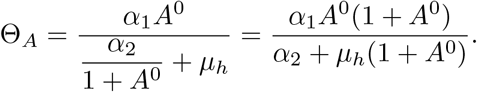

*Hence*,

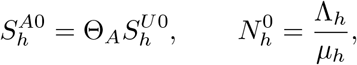

*and therefore*

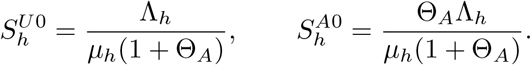

*For the mosquito population*,

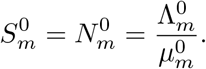

*Thus, the MFE is*

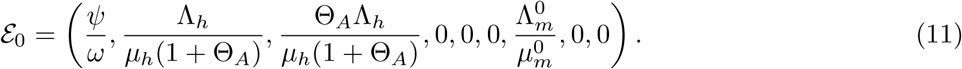

*Unlike models in which awareness vanishes in the absence of infection, the present formulation permits the coexistence of unaware and aware susceptible humans at the MFE*.

#### Definition 3

(Next-Generation Matrix Formulation). *Using the infected-state vector*

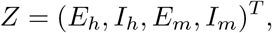

*the infection subsystem is written as*

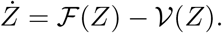

*Define the awareness-adjusted susceptible population by*

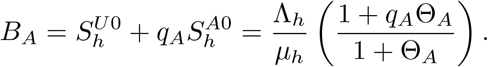

*The Jacobian matrices of new infection and transition terms, evaluated at E*_0_, *are*

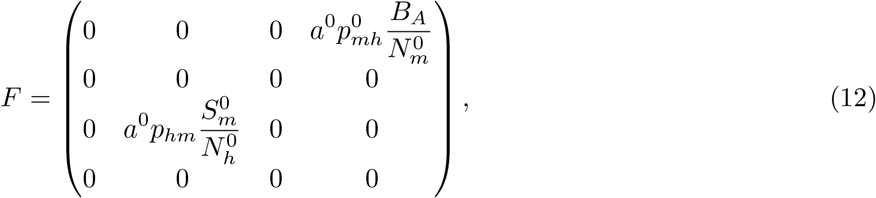

*and*

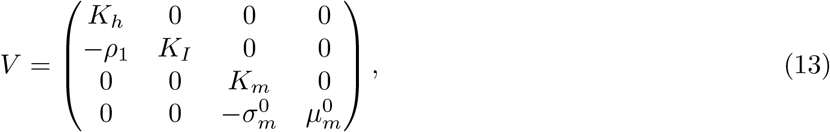

*where*

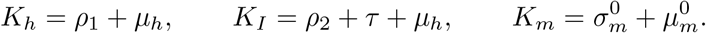

*The inverse of V is readily obtained as*

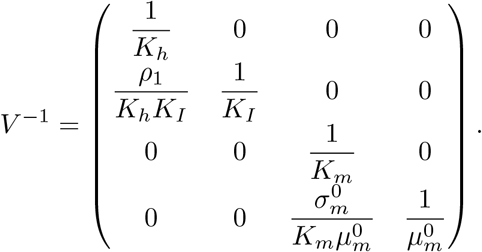

#### Definition 4

(Basic Reproduction Number). *The basic reproduction number is the spectral radius of the next-generation matrix FV* ^*−*1^:

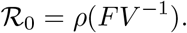

*Direct computation yields*

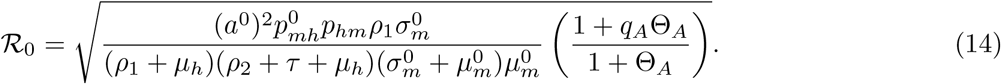

*Here*,

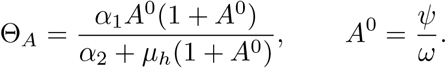

#### Remark 1

*Expression* (14) *separates the biological transmission potential from the awareness-mediated modification of human susceptibility:*

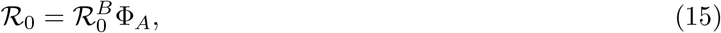

*where*

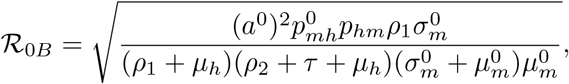

*and*

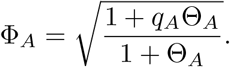

*The quantity R*_0*B*_ *represents the biological mosquito–human transmission potential under reference climatic conditions, whereas* Φ_*A*_ *is the awareness-mediated susceptibility modifier. Since* 0 *< q*_*A*_ *<* 1 *and* Θ_*A*_ *≥* 0,

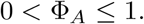

*Moreover*,

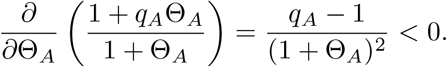

*Thus, increasing the equilibrium proportion of aware susceptible individuals reduces malaria invasion potential. This protective effect becomes stronger as q*_*A*_ *decreases*.

*The threshold interpretation is*

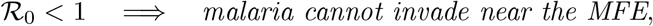

*whereas*

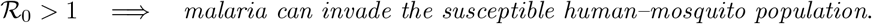

*The corresponding local stability properties are established in the following section*.

### 3.3 Local Stability of the Malaria-Free Equilibrium

The local stability of the malaria-free equilibrium *E*_0_ determines whether a small introduction of malaria infection dies out or initiates sustained transmission. The analysis is based on the frozen-climate autonomous system introduced in Section 2.

#### Theorem 4

*The malaria-free equilibrium E*_0_ *is locally asymptotically stable if R*_0_ *<* 1 *and unstable if R*_0_ *>* 1.

*Proof*. Let *J*(*E*_0_) denote the Jacobian matrix evaluated at the MFE. Using the state ordering

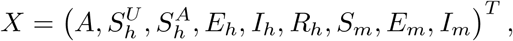

the Jacobian may be arranged in block upper-triangular form:

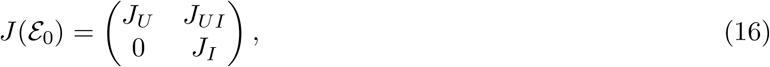

where *J*_*U*_ and *J*_*I*_ denote the uninfected and infected subsystem blocks, respectively. Hence,

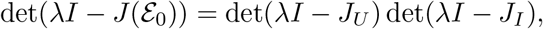

det(*λI − J*(*E*_0_)) = det(*λI − J*_*U*_) det(*λI − J*_*I*_), so the two diagonal blocks may be analysed separately.

*Uninfected subsystem*. For the awareness and susceptible-human variables 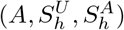, let

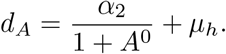

The corresponding Jacobian block is

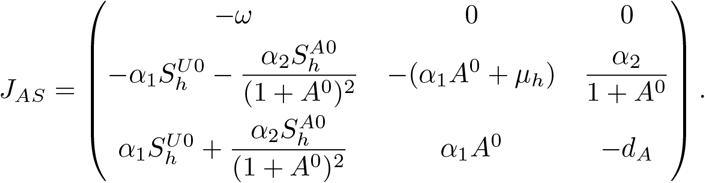

One eigenvalue is *−ω <* 0. The remaining two satisfy

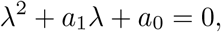

where

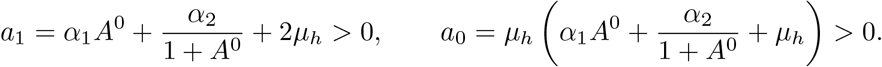

By the Routh–Hurwitz criterion, both roots have negative real parts. The recovered-human and susceptible-mosquito compartments contribute the eigenvalues

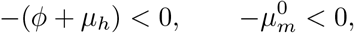

respectively. Thus, all eigenvalues associated with *J*_*U*_ have negative real parts. *Infected subsystem*. For the infected variables

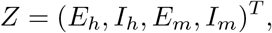

the infected Jacobian is *J*_*I*_ = *F − V*, namely

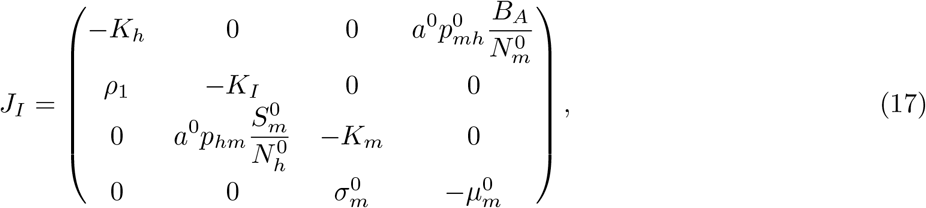

where

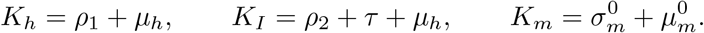

Its characteristic equation is

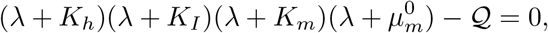

where

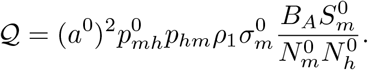

From Equation (14),

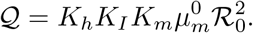

Hence, the characteristic equation becomes

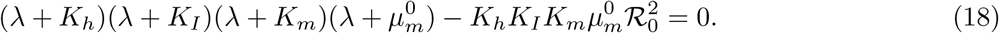

Assume, for contradiction, that there exists an eigenvalue *λ* satisfying Re(*λ*) *≥* 0. Then

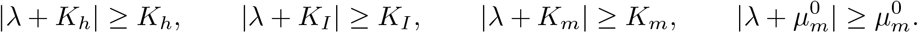

Therefore,

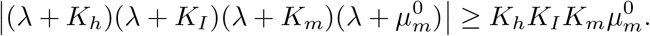

If *R*_0_ *<* 1, the right-hand constant term in Equation (18) is strictly smaller than this lower bound. Hence, the characteristic equation has no root with non-negative real part, and all eigenvalues of *J*_*I*_ lie in the open left-half plane.

Conversely, if *R*_0_ *>* 1, the characteristic polynomial *P* (*λ*) satisfies

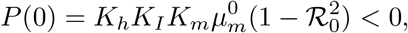

while *P* (*λ*) *→* +*∞* as *λ →* +*∞*. By continuity, *P* has at least one positive real root. Thus, *J*_*I*_ possesses a positive eigenvalue and *E*_0_ is unstable.

Therefore, *E*_0_ is locally asymptotically stable when *R*_0_ *<* 1 and unstable when *R*_0_ *>* 1.

#### Remark 2

*Biologically, Theorem 4 implies that malaria cannot invade the population when each infectious individual generates, on average, fewer than one secondary infection, that is, when R*_0_ *<* 1. *It establishes* _0_ *as the local malaria invasion threshold. Specifically*,

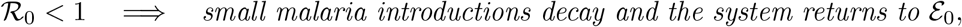

*whereas*

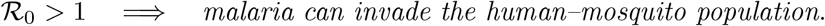

*The decomposition R*_0_ = *R*_0*B*_Φ_*A*_ *further shows that community awareness modifies the invasion threshold through*

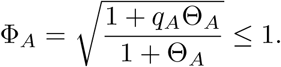

*Thus, sustained baseline awareness reduces malaria invasion potential whenever* 0 *< q*_*A*_ *<* 1. *Its protective contribution is strongest when awareness produces a substantial and persistent reduction in infection risk*.

### 3.4 Global Stability of the Malaria-Free Equilibrium

Local stability describes malaria dynamics near the malaria-free equilibrium (MFE). For elimination, however, it is necessary to determine whether infection converges to zero from arbitrary epidemiologically admissible initial conditions. We therefore establish a sufficient condition for the global asymptotic stability of _0_ in the feasible region *E* Ω, using the frozen-climate autonomous system defined in Section 2.

Recall that

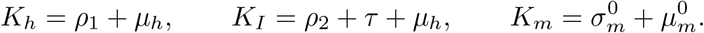

#### Remark 3

(Lyapunov Analysis). *Consider the non-negative Lyapunov function*

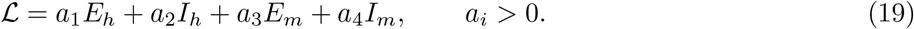

*For the autonomous uncontrolled system, the infected subsystem satisfies*

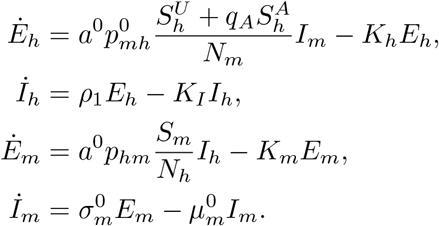

*Differentiating L and choosing*

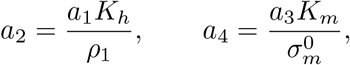

*eliminates the exposed-state terms and gives*

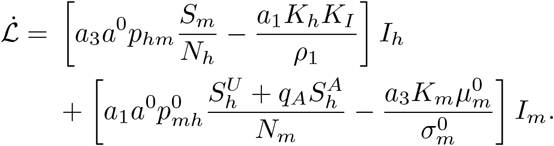

#### Definition 5

(Global Elimination Threshold). *Define the uniform transmission bounds*

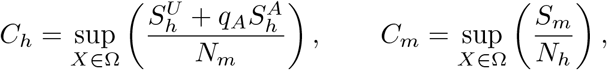

*and the global transmission threshold*

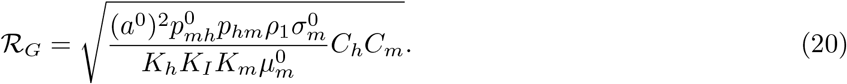

*At the MFE, the corresponding transmission ratios recover the local threshold R*_0_. *Consequently*,

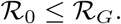

#### Theorem 5

*The malaria-free equilibrium E*_0_ *is globally asymptotically stable in* Ω *if*

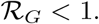

*Proof*. Using the definitions of *C*_*h*_ and *C*_*m*_,

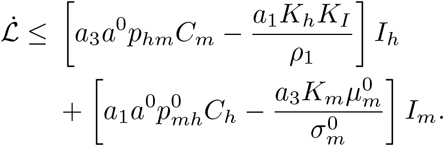

Choose *a*_1_ *>* 0 and select the constant *a*_3_ *>* 0 such that

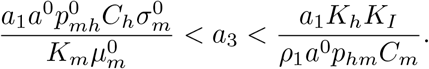

Such a choice exists precisely when *R*_*G*_ *<* 1. Hence,

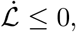

with equality only if *I*_*h*_ = *I*_*m*_ = 0. Invariance of this set further requires

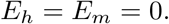

Thus, all infected compartments converge to zero.

On the infection-free invariant set, the remaining dynamics reduce to

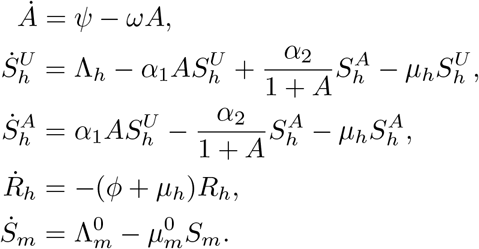

Consequently,

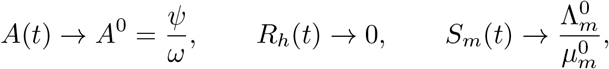

while the susceptible-human subsystem converges to

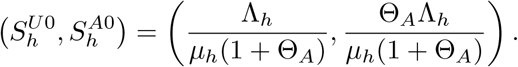

Therefore, the largest invariant subset of 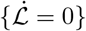 is *E*_0_ . By LaSalle’s invariance principle, *E*_0_ is globally asymptotically stable in Ω.

#### Remark 4

*Theorem 5 distinguishes the local invasion threshold R*_0_ *from the global elimination threshold R*_*G*_:

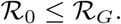

*Thus, R*_0_ *<* 1 *guarantees the decay of sufficiently small malaria introductions, whereas the stronger condition R*_*G*_ *<* 1 *ensures elimination from arbitrary admissible initial infection levels*.

*If the maximal transmission ratios are attained at the malaria-free demographic and awareness configuration, then*

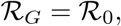

*and the classical threshold R*_0_ *<* 1 *is sufficient for global malaria elimination*.

*The distinction is particularly relevant because community awareness evolves with infection prevalence and modifies the susceptible population structure. The global threshold therefore provides a conservative elimination criterion under varying epidemiological conditions*.

### 3.5 Endemic Equilibrium and Its Existence

An endemic equilibrium represents persistent malaria transmission with positive infectious human and mosquito populations. For the frozen-climate autonomous system, let

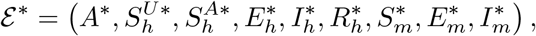

where 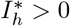 and 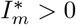. Throughout this section,

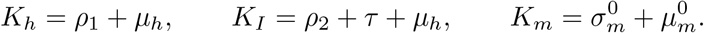

and the equilibrium forces of infection be defined by:

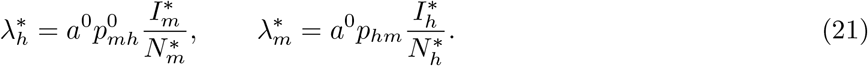

Hence,

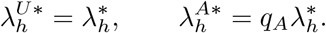

#### Definition 6

(Human Equilibrium Components). *At equilibrium, community awareness satisfies*

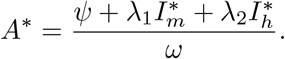

*The exposed, infectious, and recovered human populations are given by*

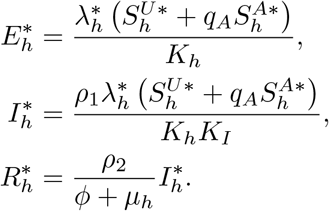

*From the aware susceptible equation*,

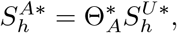

*where*

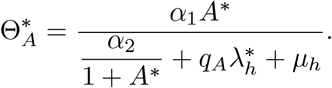

*Thus*,

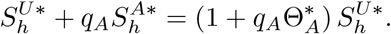

*Substitution into the unaware susceptible equilibrium equation yields*

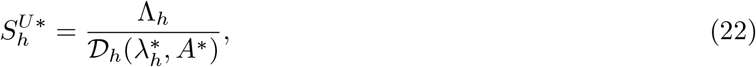

*where*

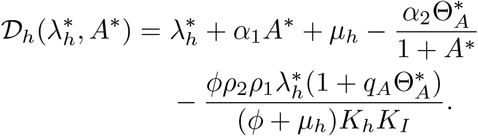

*Hence, all human equilibrium components are determined by* 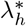 *and A*^*\**^.

#### Definition 7

(Mosquito Equilibrium Components). *The mosquito equilibrium equations yield*

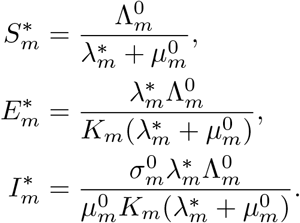

*Moreover*,

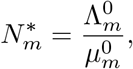

*and therefore*

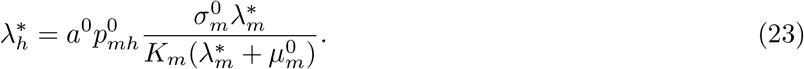

#### Definition 8

(Endemic Equilibrium Consistency Equation). *The total human population satisfies*

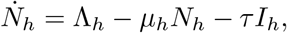

*so that, at equilibrium*,

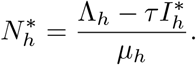

*The human-to-mosquito force of infection must also satisfy*

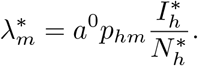

*Together with the expressions for* 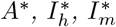, *and* 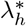, *these relations define a closed nonlinear equilibrium system. After substitution, the endemic equilibrium may be reduced to the scalar consistency equation*

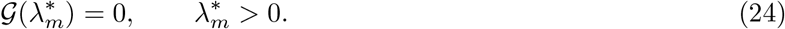

*The root* 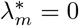 *corresponds to the MFE, whereas every positive biologically admissible root of Equation* (24) *determines an endemic equilibrium*.

#### Theorem 6

(Existence of the Endemic Equilibrium). *Suppose all model parameters are positive and the equilibrium components are biologically admissible. A branch of endemic equilibria may emerge from E*_0_ *as R*_0_ *crosses unity. If the bifurcation at R*_0_ = 1 *is forward, no nearby endemic equilibrium exists for R*_0_ *<* 1, *whereas a positive endemic equilibrium exists for R*_0_ *>* 1 *sufficiently close to unity*.

*Proof*. At 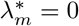, the equilibrium equations reduce to the malaria-free state *E*_0_. By Theorem 4, the MFE changes stability as *R*_0_ crosses unity. At *R*_0_ = 1, the linearized infected subsystem possesses a simple zero eigenvalue associated with the critical transmission mode.

The local emergence and direction of a non-trivial equilibrium branch are determined by the nonlinear terms of the model. A forward bifurcation yields a positive endemic branch for *R*_0_ *>* 1, whereas a backward bifurcation may permit endemic equilibria for *R*_0_ *<* 1. Accordingly, the direction of the bifurcation is established explicitly by centre manifold analysis in Section 3.6.

#### Remark 5

*The endemic equilibrium reflects a feedback between malaria prevalence and community awareness. In particular*,

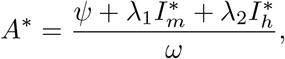

*so increasing human or mosquito infection raises the equilibrium awareness level. Greater awareness promotes movement from the unaware to the aware susceptible class, while q*_*A*_ *<* 1 *ensures that aware individuals experience lower infection risk*.

*The model therefore incorporates a negative behavioural feedback:*

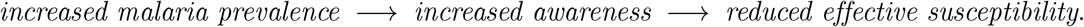

*The strength and nonlinearity of this feedback may influence both the number and stability of endemic equilibria. Consequently, the threshold R*_0_ = 1 *marks a critical transition in malaria dynamics, whose bifurcation direction is examined in the following section*.

### 3.6 Bifurcation Analysis

The threshold *R*_0_ = 1 marks a change in the local stability of the malaria-free equilibrium (MFE). However, the threshold alone does not determine whether an endemic equilibrium emerges for *R*_0_ *>* 1 or may coexist with the MFE for *R*_0_ *<* 1. Centre manifold theory is therefore used to determine the local bifurcation direction.

#### Definition 9

(Bifurcation Parameter and Centre Manifold Formulation). *The baseline mosquito-to-human transmission factor*

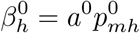

*is selected as the bifurcation parameter. From Equation* (14),

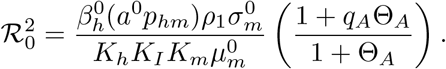

*Thus, the critical value corresponding to R*_0_ = 1 *is*

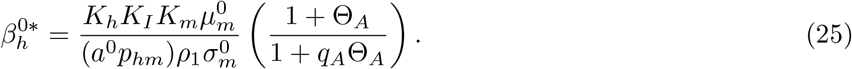

*At* 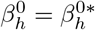, *the Jacobian at E*_0_ *possesses a simple zero eigenvalue, while the remaining eigenvalues have negative real parts*.

*Using the state ordering*

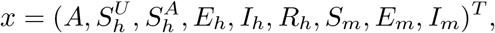

*write the system as*

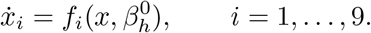

*Let w and v denote the right and left eigenvectors associated with the zero eigenvalue:*

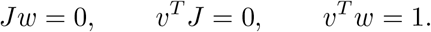

*Near* 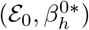, *the centre manifold dynamics reduce to*

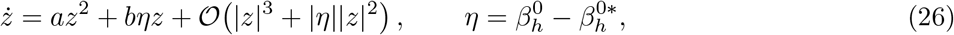

*where*

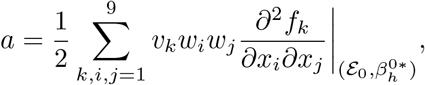

*and*

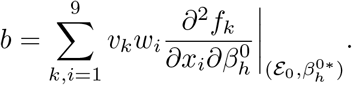

#### Remark 6

(Zero Eigenvectors). *The infected components of the right eigenvector satisfy*

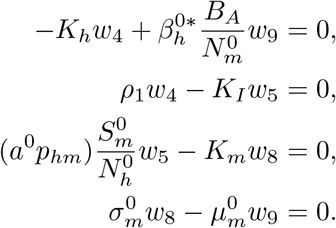

*Taking w*_9_ *>* 0 *as a scaling constant gives*

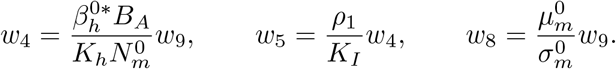

*The awareness and recovered components satisfy*

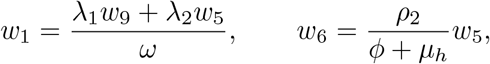

*while w*_2_, *w*_3_, *and w*_7_ *follow from the corresponding linearized demographic equations*.

*Similarly, the infected components of the left eigenvector satisfy*

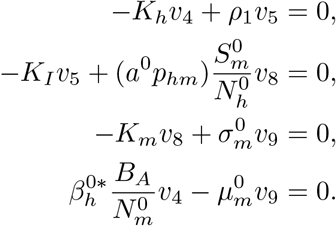

*Hence*,

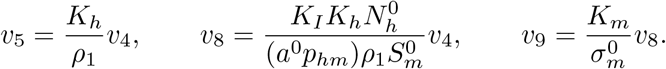

*The eigenvectors are finally rescaled so that v*^*T*^ *w* = 1.

#### Definition 10

(Bifurcation Coefficients and Direction). *The coefficient b measures the first-order effect of the bifurcation parameter on the critical transmission mode. Since* 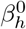 *enters the mosquito-to-human infection terms, its principal mixed derivative in the infected subsystem is*

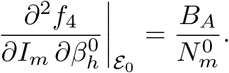

*Consequently, direct evaluation of the full centre manifold expression gives*

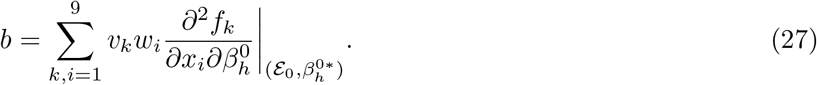

*For the baseline parameter set, the sign of b is determined numerically from this complete expression rather than from a dominant-term approximation*.

*The coefficient a incorporates nonlinear mosquito-to-human transmission, human-to-mosquito transmission, and awareness-mediated susceptibility:*

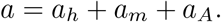

*Here, a*_*h*_ *and a*_*m*_ *represent the nonlinear transmission contributions, whereas a*_*A*_ *captures the behavioural feedback generated by*

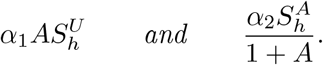

*Since*

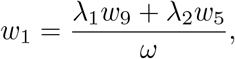

*the awareness contribution depends directly on infection-induced awareness, awareness decay, and behavioural transition rates*.

#### Theorem 7

*Suppose b >* 0 *at* 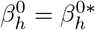. *The local bifurcation direction at R*_0_ = 1 *is determined by the sign of a:*

1. *If a <* 0, *the model undergoes a forward transcritical bifurcation, and a locally stable endemic equilibrium emerges for R*_0_ *>* 1.
2. *If a >* 0, *the bifurcation is backward, and a positive endemic equilibrium may coexist with the locally stable MFE for some R*_0_ *<* 1.

*Proof*. At *R*_0_ = 1, the Jacobian has a simple zero eigenvalue and all remaining eigenvalues have negative real parts. The centre manifold reduction therefore yields Equation (26). Under *b >* 0, the standard centre manifold criterion implies that *a <* 0 corresponds to a forward bifurcation, whereas *a >* 0 corresponds to a backward bifurcation.

#### Remark 7

*Because the coefficient a contains coupled rational incidence and awareness derivatives, its fully expanded symbolic form is cumbersome. The coefficients a and b are therefore evaluated computationally at* 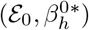 *using the baseline parameter set*.

*The numerical procedure computes the MFE and critical transmission parameter, constructs the Jacobian, obtains normalized left and right zero eigenvectors, evaluates the required second derivatives, and calculates a and b from the complete centre manifold expressions. The resulting signs determine the local bifurcation direction*.

*A bifurcation diagram is subsequently obtained by varying* 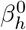 *across* 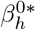 *and numerically solving for the endemic infectious-human equilibrium* 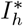.

#### Remark 8

*A forward bifurcation implies that reducing R*_0_ *below unity is locally sufficient to prevent endemic malaria persistence. A backward bifurcation, however, implies that endemic transmission may persist even when R*_0_ *<* 1.

*Dynamic awareness introduces an infection-responsive behavioural feedback: increasing malaria prevalence raises awareness, promotes protective behaviour, and reduces effective human susceptibility. Strong and persistent awareness may suppress endemic transmission, whereas rapid awareness decay or weak behavioural protection may diminish this effect. The computed signs of a and b, together with the bifurcation diagram, are reported in the numerical results*.

### 3.7 Optimal Control Problem

To assess the combined effects of malaria prevention, behavioural intervention, vector control, and case management, we consider the control vector

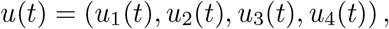

representing LLIN use, targeted awareness campaigns, indoor residual spraying (IRS), and treatment of infectious humans, respectively. The admissible control set is

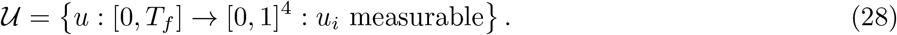

The objective is to minimize infectious humans and mosquitoes while balancing the implementation costs of the interventions. Accordingly, define

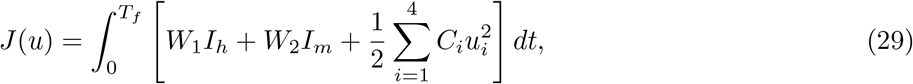

where *W*_1_, *W*_2_ *>* 0 are epidemiological weights and *C*_*i*_ *>* 0 denote intervention cost coefficients. The quadratic terms represent increasing marginal costs at higher intervention intensities.

The optimal control problem is to determine *u*^*\**^ *∈ U* such that

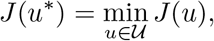

subject to system (6) and the prescribed initial conditions.

#### Theorem 8

(Existence of Optimal Controls). *There exists an optimal control u*^*\**^ *∈ U minimizing Equation* (29) *subject to the state system* (6).

*Proof*. The admissible set *U* is non-empty, closed, bounded, and convex. By Theorems 1 and 2, the state solutions remain non-negative and bounded on [0, *T*_*f*_]. The state system is continuous in the state variables and affine in the controls.

Moreover, the running cost

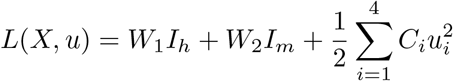

is convex in *u* and satisfies

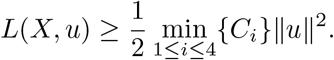

Hence, the standard existence conditions for optimal control problems are satisfied, and an optimal control *u*^*\**^ *∈ U* exists.

#### Remark 9

(Adjoint System and Optimality Conditions). *Pontryagin’s Maximum Principle is used to characterize the optimal controls. Let*

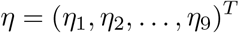

*denote the adjoint vector corresponding to*

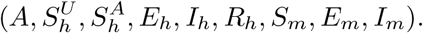

*The Hamiltonian is*

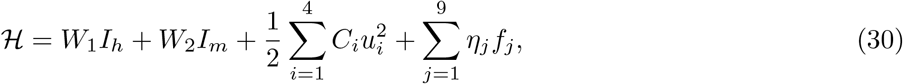

*where f*_*j*_ *denotes the right-hand side of the jth state equation*.

*The adjoint equations and transversality conditions are*

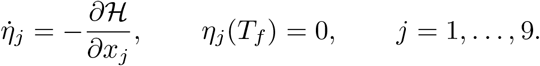

*For compactness, define*

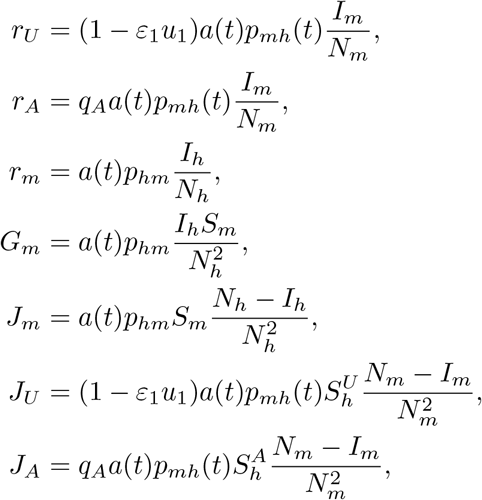

*and*

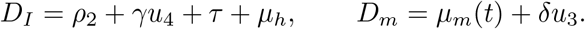

#### Remark 10

(Complete Adjoint System). *The adjoint system is*

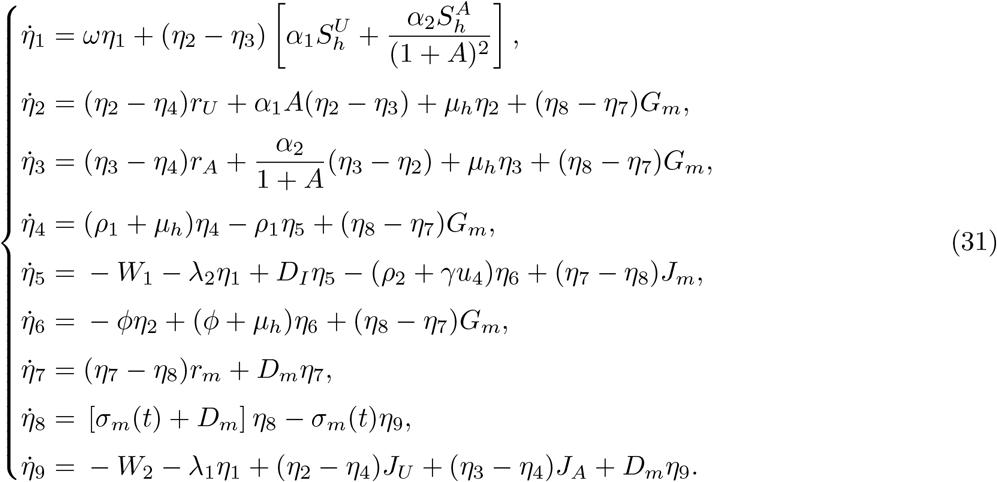

*with*

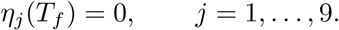

#### Definition 11

(Characterization of the Optimal Controls). *The optimal controls minimize the Hamiltonian pointwise over* [0, 1]. *Differentiating H with respect to each control gives*

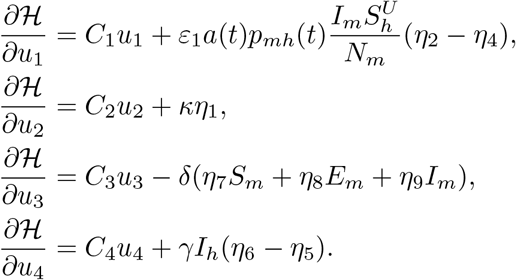

Applying the control bounds yields the following characterization.

#### Theorem 9

*Given the optimal state trajectory and corresponding adjoint variables, the optimal controls are*

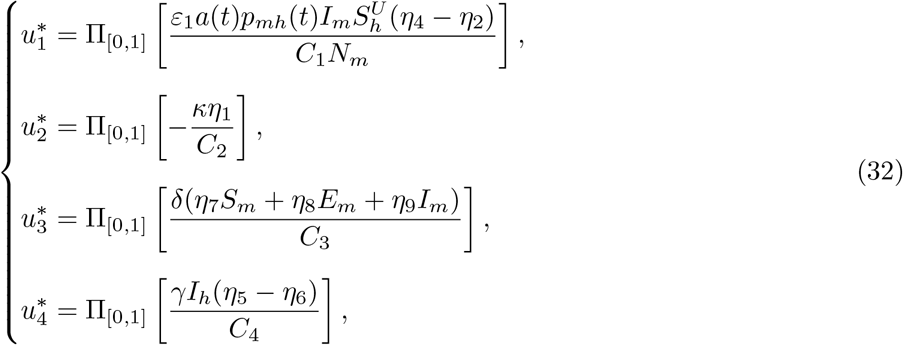

*where*

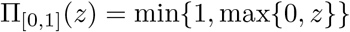

*denotes projection onto the admissible control interval*.

The optimality system therefore consists of the state equations (6), the adjoint system (31), the terminal conditions, and the control characterizations (32).

#### Remark 11

(Numerical Solution of the Optimality System). *The state-adjoint system forms a two-point boundary value problem and is solved using the forward–backward sweep method. Starting from an initial control guess, the state system is integrated forward from* 0 *to T*_*f*_ . *The adjoint system is then integrated backward from T*_*f*_ *to* 0, *and the controls are updated using Equation* (32).

*To improve numerical stability, the control update is relaxed according to*

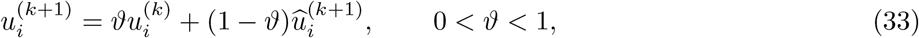

*where u*^(*k*+1)^ *is obtained directly from the optimality conditions. Iteration continues until the relative changes in the state, adjoint, and control variables fall below a prescribed tolerance*.

*The computational implementation and intervention scenarios are presented in the following section*.

### 3.8 Numerical Methods and Computational Implementation

Numerical simulations were conducted to investigate climate-driven malaria dynamics, quantify intervention effects, evaluate epidemiological thresholds, and solve the optimal control problem. All computations were implemented in MATLAB using adaptive ordinary differential equation solvers and a forward–backward sweep algorithm.

#### Numerical Integration of the State System

The nine-dimensional state system (6) was integrated over [0, *T*_*f*_] using MATLAB’s adaptive Runge–Kutta solver <monospace>ode45</monospace>. The state vector was ordered as

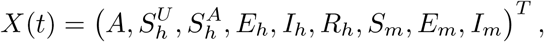

consistent with the analytical formulation and computational implementation.

Non-negative initial conditions were imposed, and the solver tolerances were specified as

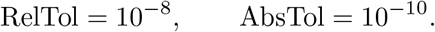

Where appropriate, the <monospace>NonNegative</monospace> solver option was imposed on all nine state variables to reduce numerical violations of biological non-negativity.

At each integration time point, the climate variables and corresponding biological response functions were evaluated before computing the state derivatives.

#### Implementation of Climate Forcing

Seasonal temperature, rainfall, and relative humidity were generated from the periodic functions defined in Equations (**??**)-(**??**). Their normalized deviations,

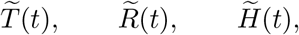

were used to update the climate-dependent biological quantities

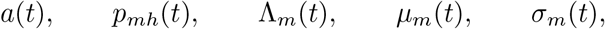

according to the response functions specified in the model formulation.

Thus, the numerical algorithm followed the sequence

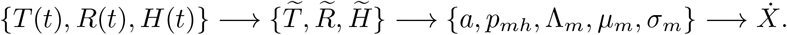

The modular implementation also permits the periodic climate functions to be replaced by interpolated meteorological observations without modifying the epidemiological state equations.

#### Computation of *R*_0_ and Sensitivity Indices

Under reference climatic conditions, the basic reproduction number was evaluated from

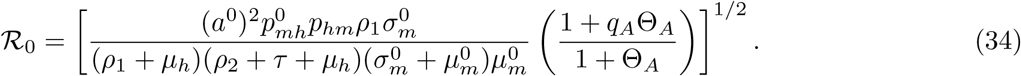

Normalized forward sensitivity indices were computed as

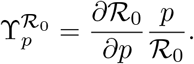

Closed-form expressions derived in Section **??** were used whenever available. Numerical verification was performed using the centred finite-difference approximation

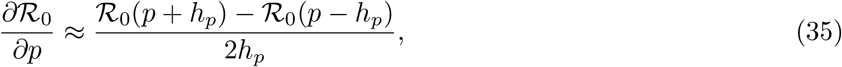

where *h*_*p*_ was selected relative to the magnitude of *p*. Sensitivity indices were ranked according to 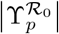.

#### Bifurcation Computation

The critical mosquito-to-human transmission factor 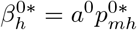 was obtained by imposing *R*_0_ = 1. At the critical value, the Jacobian matrix was evaluated at the malaria-free equilibrium and the eigenvalue of smallest absolute magnitude was identified.

The associated right and left eigenvectors, *w* and *v*, were computed numerically and normalized such that

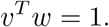

The centre manifold coefficients *a* and *b* were then evaluated from the complete expressions given in Section 3.6. Required second-order derivatives of the vector field were approximated numerically using centred finite differences.

The sign of *b* was first verified computationally. For *b >* 0, the sign of *a* determined the local bifurcation direction: *a <* 0 indicated a forward bifurcation, whereas *a >* 0 indicated a backward bifurcation.

To construct the bifurcation diagram, the transmission parameter was varied across its critical value and the nonlinear equilibrium equations were solved numerically. The endemic infectious-human population 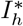 was used as the bifurcation response variable.

#### Forward–Backward Sweep Algorithm

The optimality system was solved using the forward–backward sweep method on the time grid

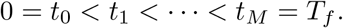

##### Algorithm 1

Forward–Backward Sweep Method

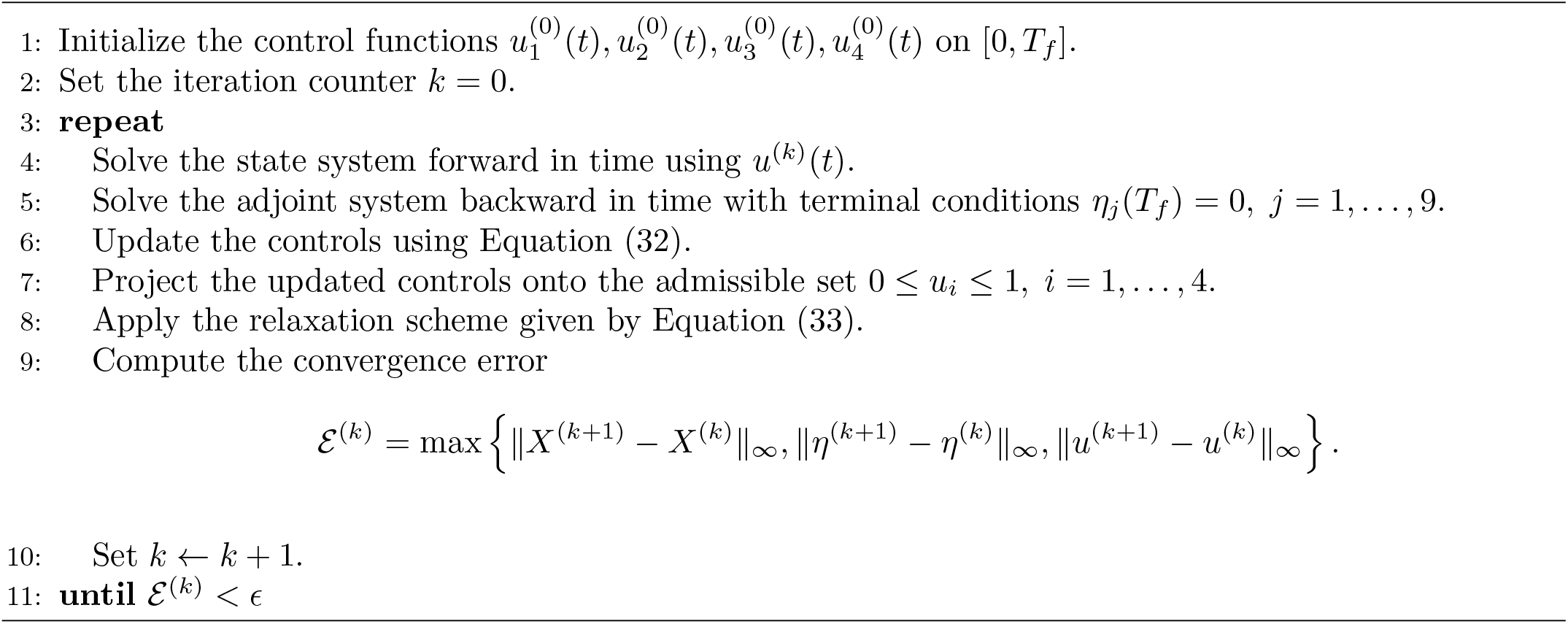

#### Intervention Scenarios and Outcome Measures

The uncontrolled baseline was compared with selected single and combined intervention strategies:

*S*_0_ : no intervention,

*S*_1_ : LLINs only,

*S*_2_ : awareness campaigns only,

*S*_3_ : IRS only,

*S*_4_ : treatment only,

*S*_5_ : LLINs and awareness,

*S*_6_ : LLINs, IRS, and treatment,

*S*_7_ : all four interventions.

For each strategy, the cumulative infectious-human burden was approximated by

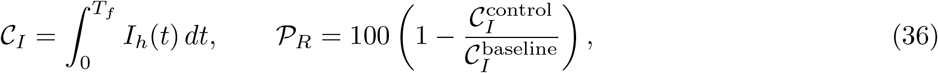

where *P*_*R*_ denotes the percentage reduction relative to the uncontrolled baseline.

The principal numerical outcomes were the peak infectious-human population, time to peak infection, cumulative infectious-human burden, percentage reduction in disease burden, objective functional value, and optimal intervention profiles. These measures were used to compare the epidemiological performance of the alternative intervention strategies.

## 4 Numerical Results

Numerical simulations were conducted to investigate the temporal behaviour of the climate-driven SEAIR–SEI malaria model, quantify the epidemiological effects of intervention coverage, evaluate the response of malaria transmission to alternative climate scenarios, and identify the parameters that exert the greatest influence on disease burden. Unless otherwise specified, simulations were performed over a 365-day period with initial human and mosquito populations of 139,970 and 500,600, respectively. The reference environmental conditions were a mean temperature of 25°C, rainfall of 120 mm/month, and relative humidity of 75%. The baseline mosquito biting rate was *a* = 0.33 bites per mosquito per day, while the mosquito-to-human and human-to-mosquito transmission probabilities were *p*_*mh*_ = 0.20 and *p*_*hm*_ = 0.50, respectively.

### 4.1 Baseline malaria transmission dynamics

The baseline simulation revealed a distinct seasonal malaria outbreak, followed by a gradual decline in infection prevalence. The infectious human population attained a maximum 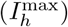 of 193.99 on day 110.79. The infectious mosquito population reached a substantially larger numerical peak 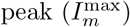 of 1272.59 on day 122.72. Thus, the mosquito infection peak occurred approximately 11.93 days after the peak in human infection. The temporal ordering of the two peaks reflects the coupled host-vector feedback mechanism represented in the model. Increased infectious human prevalence raises the force of infection experienced by susceptible mosquitoes. Following the mosquito latent period, the resulting increase in infectious vectors sustains mosquito-to-human transmission. The relatively short separation between the human and mosquito peaks indicates strong epidemiological coupling between the two infectious populations.

At the end of the simulation period, the infectious human population had declined to 5.16 individuals, whereas the exposed human population was 3.48. The recovered population reached 804.82. A pronounced redistribution of the susceptible human population was observed between the awareness classes: 136,175.46 individuals were in the aware susceptible class, compared with 2,978.65 in the unaware susceptible class. The awareness variable reached 107.02 at the end of the simulation.

The cumulative human infection burden, measured as the area under the infectious-human trajectory, 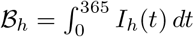 *dt* was estimated at 40,039.25 person-days. Unlike peak prevalence, this quantity captures both the magnitude and duration of infectious human prevalence and therefore provides a more comprehensive measure of epidemic burden.

Figure 2 illustrates the baseline infection trajectories. The results demonstrate that reliance on the peak number of infectious individuals alone may provide an incomplete description of the epidemic. Although the infectious populations eventually declined to low levels, the cumulative burden remained substantial because infection persisted over an extended portion of the simulation horizon.

**Fig 2.**
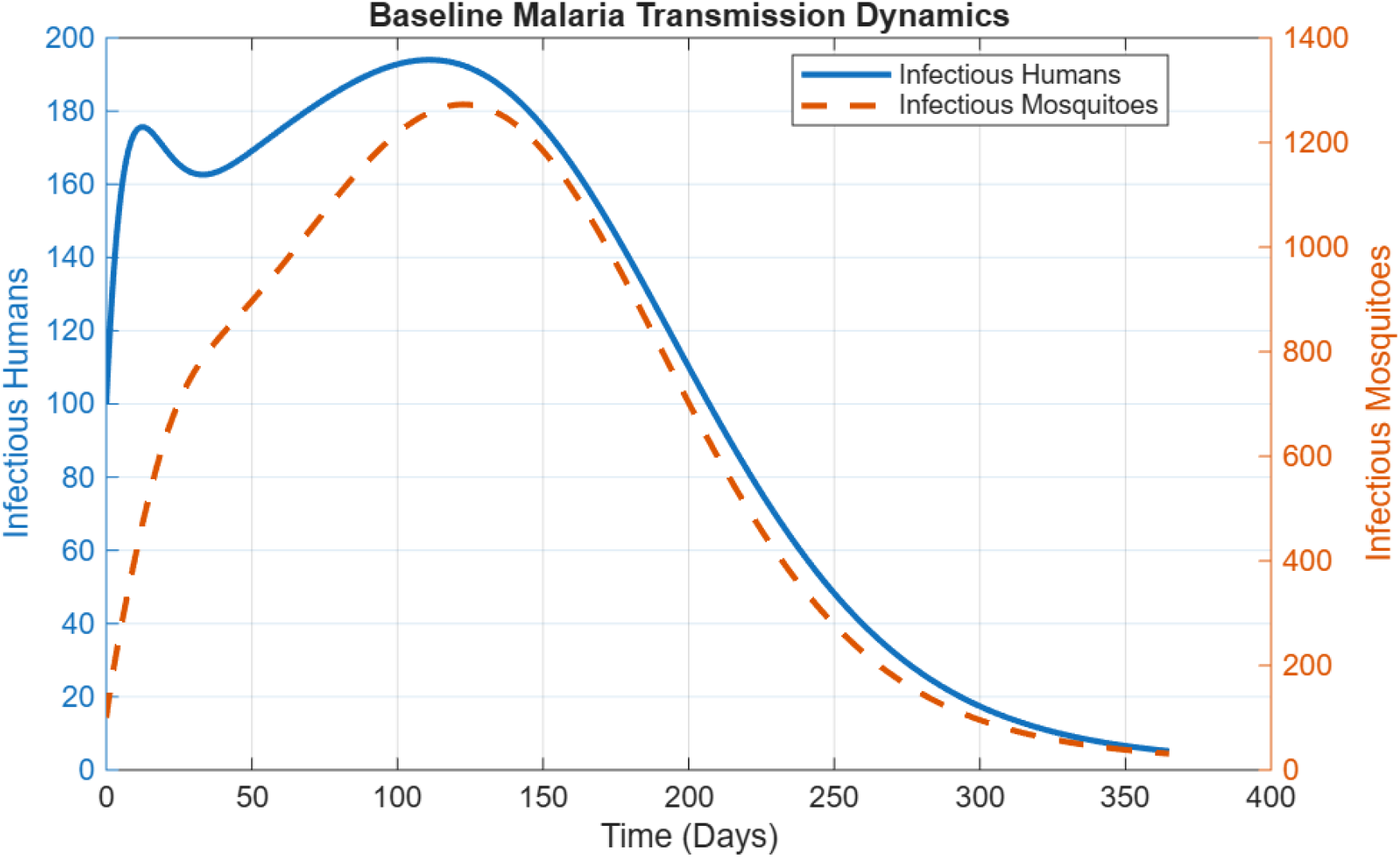
Baseline dynamics of infectious humans and infectious mosquitoes over the 365-day simulation period.

### 4.2 Effects of intervention coverage

The impact of long-lasting insecticide treated bed nets (LLINs), community awareness, indoor residual spraying (IRS), prompt treatment, and a strategy comprising more than one intervention were assessed at 20%, 40%, 60%, 80%, and 100% coverage levels. The effectiveness of control measures was evaluated using two alternative epidemiological endpoints: the reduction of peak human infectious prevalence, and the reduction of total human infection burden.

Table 3 and Figure 3(a) show that increasing intervention coverage consistently reduced the epidemic peak, although the effectiveness varied substantially among strategies. Prompt treatment produced the greatest reduction in peak infectious prevalence across all coverage levels, lowering the epidemic peak by nearly one-half (47.53%) at full coverage. The combined strategy achieved only a marginally larger reduction (47.57%), whereas LLINs, IRS, and awareness campaigns produced comparatively modest reductions of 13.14%, 10.15%, and 9.48%, respectively.

**Table 2.**
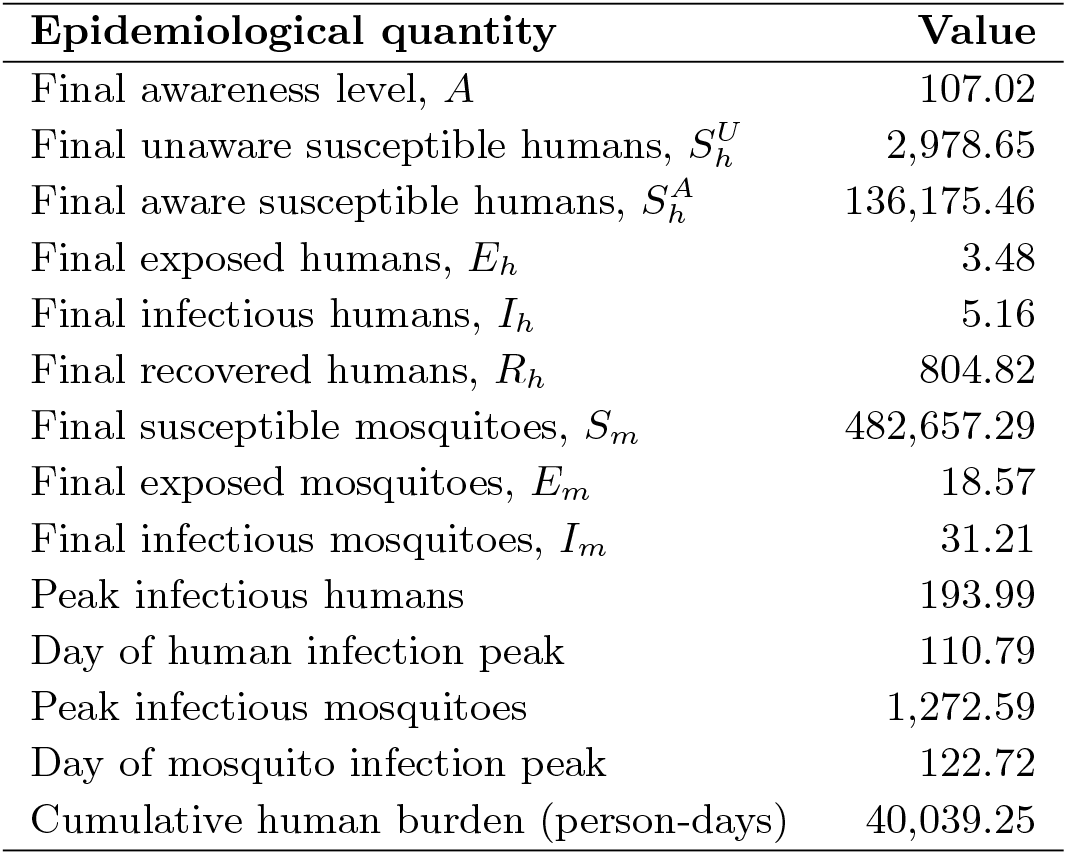
Summary of baseline malaria dynamics over the 365-day simulation period.

**Table 3.** Percentage reduction in peak infectious human prevalence under alternative intervention coverage levels.

| Coverage (%) | LLINs | Awareness | IRS | Treatment | Combined |
| --- | --- | --- | --- | --- | --- |
| 20 | 10.34 | 5.18 | 9.61 | 24.82 | 25.20 |
| 40 | 11.13 | 9.01 | 9.75 | 34.26 | 34.61 |
| 60 | 11.85 | 9.47 | 9.89 | 40.59 | 40.84 |
| 80 | 12.52 | 9.48 | 10.02 | 44.88 | 45.02 |
| 100 | 13.14 | 9.48 | 10.15 | 47.53 | 47.57 |

**Fig 3.**
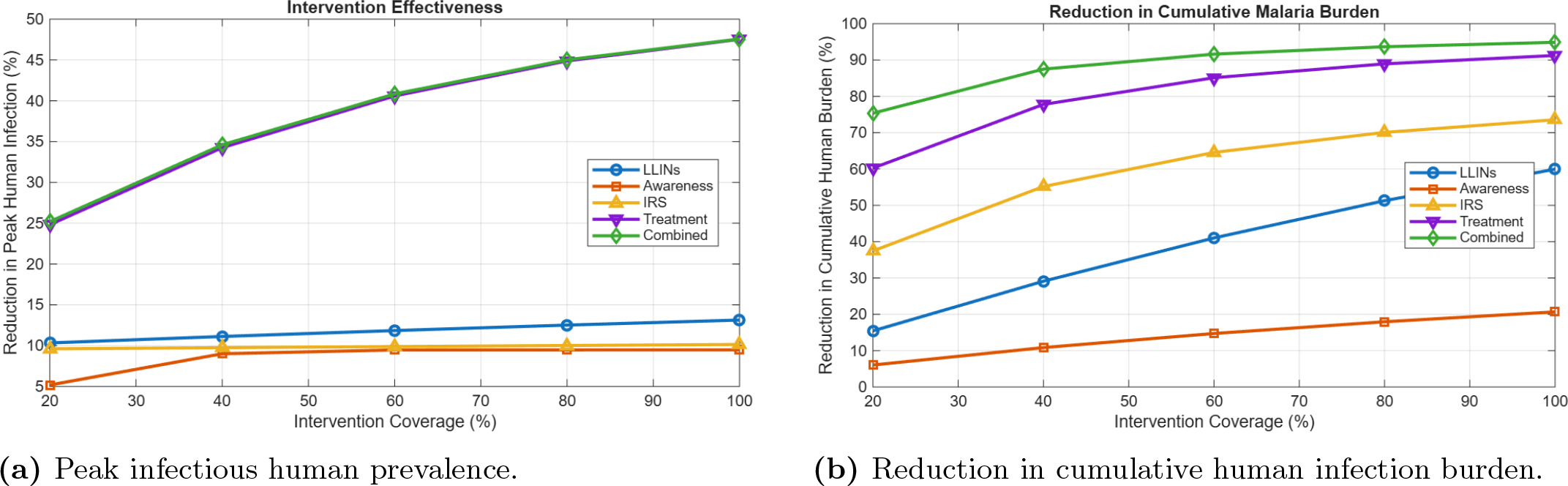
Effects of intervention coverage on malaria dynamics. (a) Reduction in peak infectious human prevalence under increasing intervention coverage. (b) Percentage reduction in cumulative human infection burden for individual and combined intervention strategies.

The relatively small difference between prompt treatment and the combined strategy indicates that treatment primarily determines the instantaneous epidemic peak because it shortens the infectious period. Preventive interventions such as LLINs, IRS, and awareness campaigns instead reduce onward transmission, and their greatest epidemiological benefits are therefore expected to emerge over the duration of the epidemic rather than at its peak.

Table 4 together with Figure 3(b) indicates a different story for cumulative disease burden. Combined intervention was generally more effective than any single intervention strategy as it reduced the cumulative human infection by 75.35% and 94.87% at 20% and 100% coverage respectively. Indeed, prompt treatment strategy also yielded a significant decrease in disease burden (91.25%). However, the combined strategy of prompt treatment with the use of LLINs, IRS and awareness campaigns had an additional benefit of reducing the overall disease transmission. In terms of individual preventive interventions, IRS was found to be the most efficient, followed by LLINs and lastly awareness campaigns which had the least independent effect on reducing the disease burden at all coverage levels.

**Table 4.** Percentage reduction in cumulative human infection burden under alternative intervention coverage levels.

| Coverage (%) | LLINs | Awareness | IRS | Treatment | Combined |
| --- | --- | --- | --- | --- | --- |
| 20 | 15.45 | 6.05 | 37.45 | 60.25 | 75.35 |
| 40 | 29.11 | 10.84 | 55.18 | 77.78 | 87.50 |
| 60 | 41.03 | 14.73 | 64.58 | 85.12 | 91.62 |
| 80 | 51.28 | 17.95 | 70.07 | 88.95 | 93.65 |
| 100 | 59.95 | 20.67 | 73.57 | 91.25 | 94.87 |

**Table 5.** Epidemiological outcomes under alternative climate scenarios.

| Scenario | Peak $I_h$ | $\Delta I_h$ (%) | Peak $I_m$ | $\Delta I_m$ (%) | Burden | $\Delta B_h$ (%) |
| --- | --- | --- | --- | --- | --- | --- |
| Baseline | 193.99 | 0.00 | 1,272.59 | 0.00 | 40,039 | 0.00 |
| +1°C | 264.32 | 36.25 | 1,831.60 | 43.93 | 54,091 | 35.10 |
| +2°C | 374.31 | 92.95 | 2,719.30 | 113.68 | 75,675 | 89.00 |
| Rainfall +20% | 194.12 | 0.07 | 1,274.40 | 0.14 | 40,066 | 0.07 |
| Rainfall -20% | 193.87 | -0.07 | 1,270.80 | -0.14 | 40,012 | -0.07 |
| Humidity +10% | 208.23 | 7.34 | 1,455.70 | 14.39 | 43,156 | 7.78 |
| Humidity -10% | 181.24 | -6.58 | 1,113.50 | -12.50 | 37,186 | -7.13 |
| +2°C, rainfall +20% | 374.64 | 93.12 | 2,723.60 | 114.02 | 75,741 | 89.17 |
| +2°C, rainfall -20% | 373.98 | 92.78 | 2,715.00 | 113.34 | 75,609 | 88.84 |
| +2°C, rainfall +20%, humidity +10% | 408.01 | 110.32 | 3,147.10 | 147.30 | 82,873 | 106.98 |

Figure 3 shows a descriptive summary of the intervention performance. First, panel (a) suggests that while the reduction in peak prevalence increases rapidly with coverage, it saturates at higher coverage levels for all interventions, with LLINs, IRS, and awareness campaigns having the strongest impact. Similar saturation effects have been reported in previous malaria modeling studies, where increasing intervention coverage beyond certain thresholds produces diminishing epidemiological returns because of nonlinear transmission dynamics [3, 6, 7]. Second, panel (b) reveals that the cumulative disease burden decreases steadily with coverage for each intervention, consistent with previous simulation studies demonstrating that sustained increases in intervention coverage reduce malaria burden by suppressing both human and vector infection reservoirs [4, 15]. Most importantly, the combined strategy dominates all other interventions at all coverage levels. This finding agrees with previous mathematical modeling studies and systems-based reviews showing that integrated malaria control strategies outperform individual interventions because they simultaneously target multiple components of the transmission cycle, thereby producing synergistic reductions in disease burden [3, 6, 14]. While the rapid reduction in peak prevalence suggests that timely treatment is the most efficient measure to lower the height of the epidemic peak, our results highlight the importance of adopting a combination of measures to drastically reduce the overall force of infection by concurrently lowering infectiousness, contact rates, vector densities, and human biting rates. Similar conclusions have been drawn by [14], who argued that the complexity of malaria transmission necessitates multidisciplinary and integrated intervention strategies, and by the WHO, which recommends combining vector control, case management, surveillance, and community engagement for sustainable malaria control [24]. Overall, the results suggest that coordinated multi-intervention strategies are more effective in reducing the burden of malaria than relying on a single intervention, corroborating previous mathematical modeling studies and systems-based analyses of malaria control [6, 14, 24].

### 4.3 Climate-driven changes in malaria transmission

The expanded climate scenario analysis examined independent and combined perturbations in temperature, rainfall, and relative humidity. In addition to epidemiological outcomes, the simulations tracked climate-dependent changes in mosquito biting rate, mosquito-to-human transmission probability, mosquito recruitment, mosquito mortality, and the extrinsic incubation rate.

Temperature turned out to be the most influential factor in the scenario analysis. Elevating the mean temperature by 1°C raised the number of peak infected humans by 36.25%, peak infected mosquitoes by 43.93%, and the total number of infected humans by 35.1%. By increasing the temperature by 2°C, the peak of human infections rose by 92.95%, and the peak of mosquito infections jumped by 113.68%. Meanwhile, the aggregate number of infected people climbed by 89

Temperature stress resulted in higher mean biting rate, which grew from 0.33119 to 0.35877. Likewise, the mean transmission probability from mosquitoes to humans increased from 0.20030 to 0.21056, while the average extrinsic incubation period rose from 0.10081 to 0.11366. Thus, it is evident that warming led to greater disease persistence and prevalence due to changes in more than one transmission parameter, as opposed to a change in a single climate-dependent parameter. Compared to temperature, the impact of rainfall perturbations was relatively limited under the chosen parameterization. A 20% rise in rainfall resulted in a meager 0.07% increase in peak infected humans and a 0.07% rise in overall disease burden. Meanwhile, a 20% decline in rainfall resulted in almost identical (yet negative) changes in these outcome variables. Thus, it may be concluded that, under the given set of response functions and baseline environmental conditions, the impact of rainfall on mosquito recruitment was lower than the effect of temperature on biting and pathogen incubation. Finally, the influence of humidity was moderate compared to temperature but stronger than that of rainfall. The 10% rise caused a 7.34% increase in the number of peak infected humans, a 14.39% rise in peak infected mosquitoes, and a 7.78% climb in total infected humans. Meanwhile, the analogous decrease in humidity led to a 6.58% decrease in peak infected humans, a 12.5% drop in peak infected mosquitoes, and a 7.13% reduction in the aggregate number of infected humans. The drop in humidity led to a rise in mean mosquito mortality from 0.06935 to 0.07363. The scenario that combined +2°C warming, 20% increase in rainfall, and a 10% rise in humidity should be selected if no changes in the environment are desirable. Under this scenario, the peak number of infectious humans equals 408.01, which is 136.252% of the baseline scenario (Figures 4 and 5). The peak number of infected mosquitoes is 3,147.10, an increase of 147.30%. Finally, the aggregate number of infected humans is 82,872.59 person-days, which is 106.98% above the baseline level.

**Fig 4.**
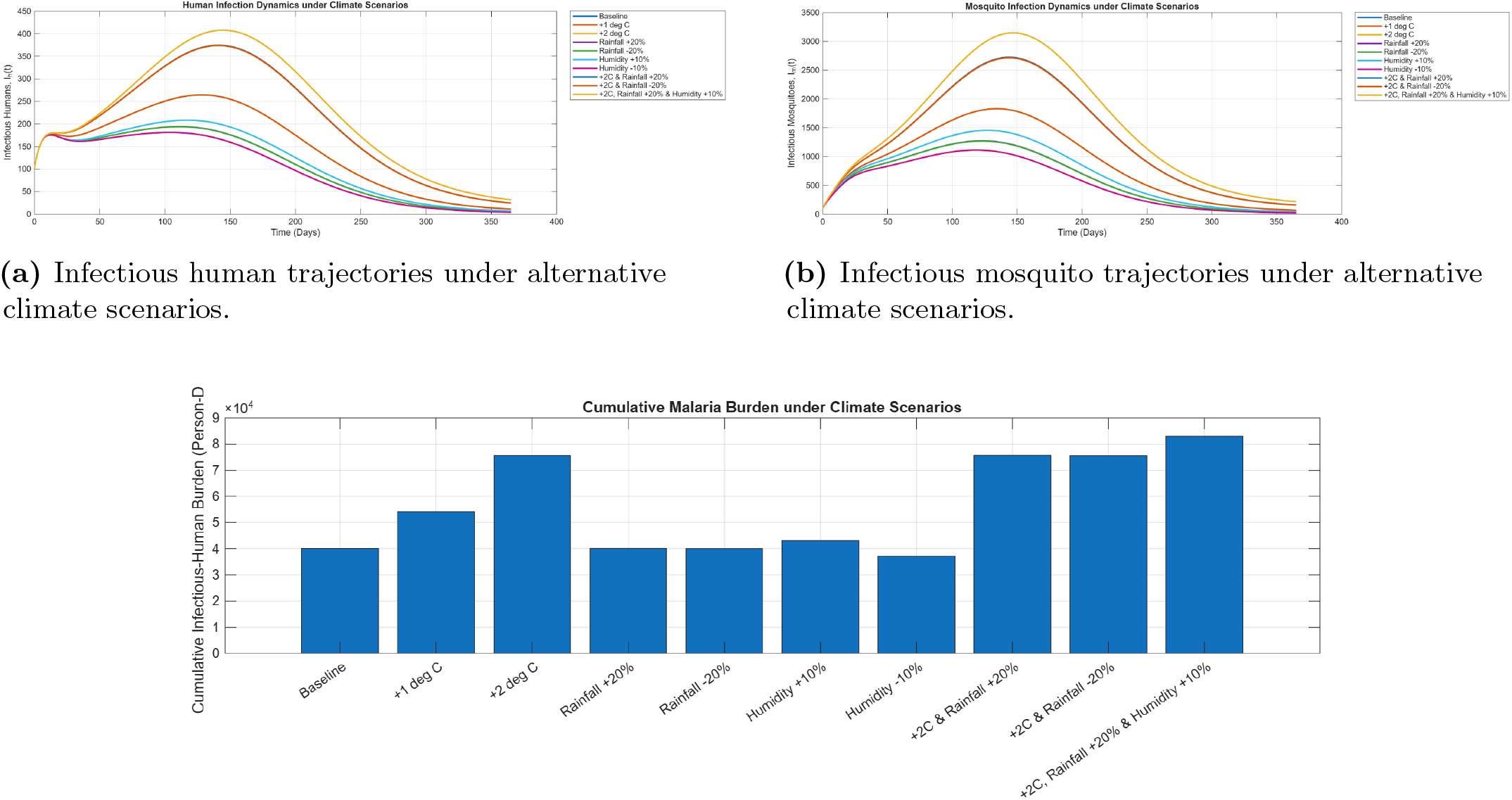
Effects of alternative climatic scenarios on malaria transmission dynamics. (a) Infectious human trajectories under different temperature, rainfall, and relative humidity conditions. (b) Corresponding infectious mosquito trajectories. (c) Cumulative human malaria burden under the alternative climate scenarios.

**Fig 5.**
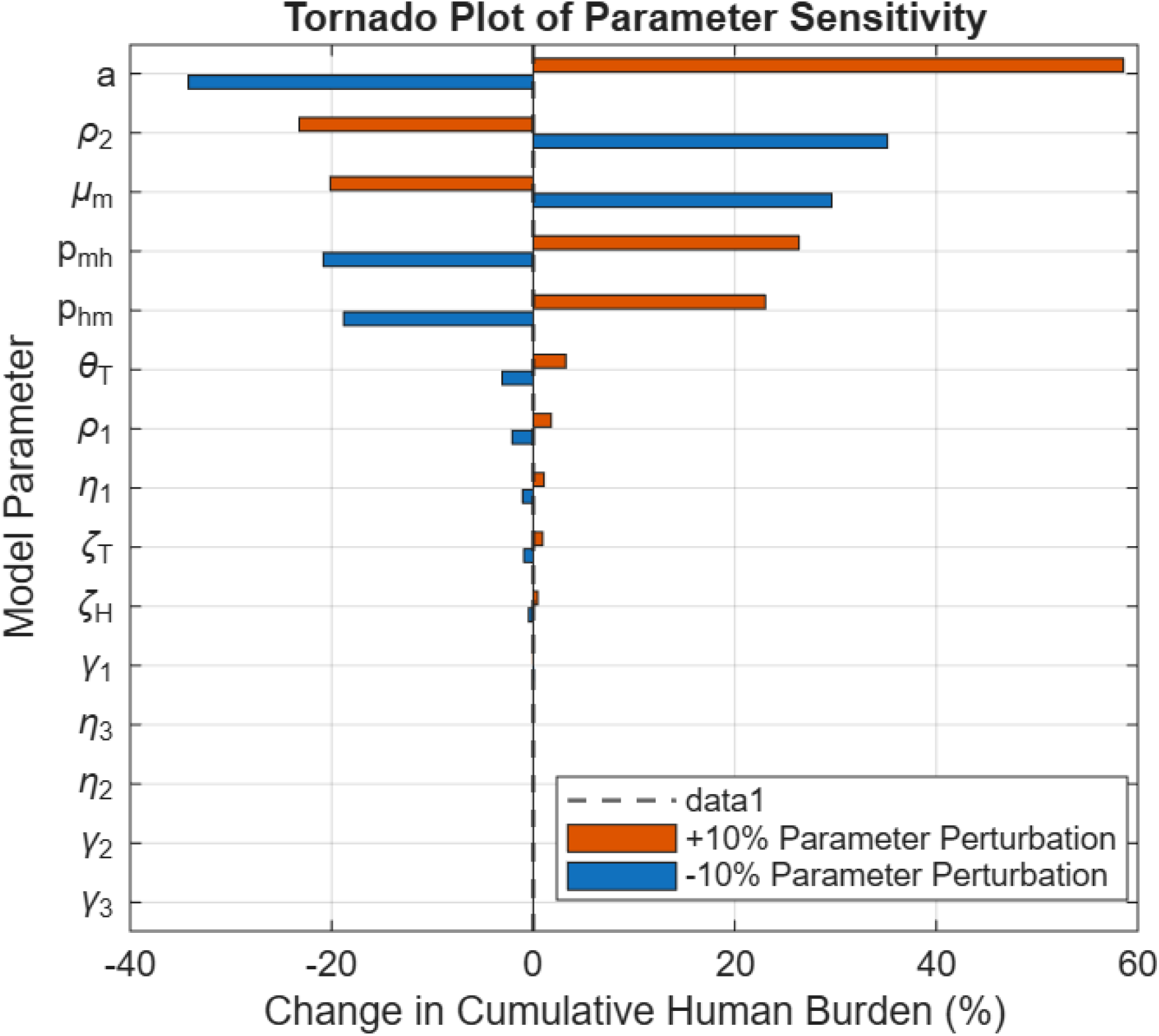
Tornado plot showing the effects of *±* 10% perturbations in selected model parameters on cumulative human malaria burden. Parameters are ordered according to the magnitude of their normalized elasticity.

The climate results demonstrate that environmental changes can influence malaria burden through several mutually reinforcing biological pathways. In particular, temperature simultaneously altered mosquito biting, transmission efficiency, recruitment, and parasite development, consistent with previous studies showing that climatic variables regulate multiple components of the malaria transmission cycle rather than acting through a single mechanism [13, 17]. The resulting epidemiological response was therefore strongly nonlinear when multiple favourable climate conditions occurred concurrently, corroborating mathematical and empirical studies demonstrating that interactions among temperature, rainfall, humidity, vector ecology, and parasite development can amplify malaria transmission under suitable environmental conditions [1, 4, 6]. These findings further support the systems perspective that malaria transmission emerges from complex interactions among climatic, biological, and behavioural processes, making integrated climate-sensitive control strategies essential for sustainable malaria management [14, 24].

### 4.4 Sensitivity and Elasticity Analysis

A normalized one-at-a-time sensitivity analysis was performed by applying ±10% perturbations to selected model parameters around their baseline values. The effects of these perturbations were evaluated with respect to peak human infection, peak mosquito infection, and cumulative human infection burden. Because the model parameters differ in scale and units, normalized elasticities were used to facilitate direct comparison of their relative epidemiological importance.

The normalized elasticity of cumulative human infection burden with respect to parameter *p* was approximated by

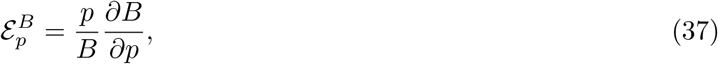

where *B* denotes the cumulative human infection burden. Positive elasticities indicate that increasing the parameter increases malaria burden, whereas negative elasticities indicate that increasing the parameter reduces disease burden.

The resulting elasticity ranking is presented in Table 6, while the corresponding tornado plot is shown in Figure 5.

**Table 6.** Ranking of model parameters according to the absolute normalized elasticity of cumulative human infection burden.

| Rank | Parameter | Symbol | Burden elasticity |
| --- | --- | --- | --- |
| 1 | Reference mosquito biting rate | $a^0$ | 4.6397 |
| 2 | Human recovery rate | $\rho_2$ | -2.9186 |
| 3 | Reference mosquito mortality rate | $\mu_m^0$ | -2.4878 |
| 4 | Reference mosquito-to-human transmission probability | $p_{mh}^0$ | 2.3597 |
| 5 | Human-to-mosquito transmission probability | $p_{hm}$ | 2.0921 |
| 6 | Temperature sensitivity of biting rate | $\theta_T$ | 0.3158 |
| 7 | Human latent progression rate | $\rho_1$ | 0.1937 |
| 8 | Temperature sensitivity of $p_{mh}(t)$ | $\chi_{pT}$ | 0.1057 |
| 9 | Temperature sensitivity of $\sigma_m(t)$ | $\chi_{\sigma T}$ | 0.0933 |
| 10 | Humidity sensitivity of $\sigma_m(t)$ | $\chi_{\sigma H}$ | 0.0453 |
| 11 | Temperature sensitivity of $\Lambda_m(t)$ | $\chi_{\Lambda T}$ | -0.0121 |
| 12 | Humidity sensitivity of $p_{mh}(t)$ | $\chi_{pH}$ | 0.0016 |
| 13 | Rainfall sensitivity of $p_{mh}(t)$ | $\chi_{pR}$ | 0.0006 |
| 14 | Rainfall sensitivity of $\Lambda_m(t)$ | $\chi_{\Lambda R}$ | -0.0003 |
| 15 | Humidity sensitivity of $\Lambda_m(t)$ | $\chi_{\Lambda H}$ | -0.0002 |

Figure 5 confirms the ranking presented in Table 6. The reference mosquito biting rate, *a*^0^, was the dominant determinant of malaria burden, with a normalized elasticity of 4.6397. Consequently, a 1% proportional increase in the biting rate produced an approximately 4.64% increase in cumulative human infection burden. The corresponding elasticities for peak human and peak mosquito infections were 3.5650 and 5.3390, respectively. Similar findings have been reported in several malaria sensitivity analyses, where the mosquito biting rate consistently emerged as one of the most influential parameters governing malaria transmission because it directly determines the frequency of vector–host contacts and the force of infection [7, 16, 22].

The human recovery rate *ρ*_2_ ranked second, with a negative elasticity of *−*2.9186, indicating that faster recovery substantially reduced disease burden. Similarly, the reference mosquito mortality rate 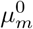 had an elasticity of *−*2.4878, demonstrating the importance of interventions that reduce adult mosquito survival. These findings agree with comparative sensitivity analyses showing that mosquito mortality and human recovery are among the principal determinants of malaria prevalence and therefore represent key targets for vector control and effective case management [7, 22, 24].

The reference mosquito-to-human transmission probability 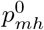 and the human-to-mosquito transmission probability *p*_*hm*_ also exhibited large positive elasticities. Together with the dominant influence of the biting rate, these findings indicate that the vector–host contact process is the primary driver of malaria transmission in the model. This observation is consistent with both classical Ross–Macdonald theory and modern benchmark studies comparing multiple malaria transmission models [11, 18, 20, 22].

Among the climate-response parameters, the temperature sensitivity of the biting rate, *θ*_*T*_, had the largest influence on cumulative malaria burden, followed by the temperature sensitivities of mosquito-to-human transmission (*χ*_*pT*_) and the mosquito extrinsic incubation rate (*χ*_*σT*_). In contrast, the rainfall-dependent response coefficients exhibited comparatively small elasticities under the baseline parameterization. These findings are consistent with mechanistic and experimental studies demonstrating that temperature simultaneously regulates mosquito biting frequency, adult survival, parasite development, and vector competence, thereby exerting a stronger overall influence on malaria transmission than rainfall acting through breeding habitat alone [1, 4, 13, 17, 19].

Overall, the sensitivity analysis demonstrates that biological transmission parameters governing mosquito biting, transmission efficiency, mosquito survival, and human recovery dominate malaria dynamics, whereas the direct effects of climatic response coefficients are comparatively modest. This finding explains why temperature exerts a stronger epidemiological influence than rainfall, as temperature simultaneously modifies several biological processes to which disease burden is intrinsically highly sensitive. Similar conclusions have been drawn by [22], who showed that despite structural differences among malaria models, the dominant sensitivity parameters are consistently associated with mosquito biting, mosquito survival, and human recovery, and by [19], who experimentally demonstrated that temperature jointly influences these same biological processes.

### 4.5 Reproduction Number, Threshold Verification, and Bifurcation Behaviour

The basic reproduction number was evaluated for the uncontrolled system using the next-generation matrix (NGM) approach under frozen reference climatic conditions. At these conditions, the mosquito biting rate was *a* = 0.33, the mosquito-to-human transmission probability was 0.19979, the mosquito mortality rate was 0.07240, and the extrinsic incubation rate was 0.10000.

Prior to constructing the NGM, the MFE was numerically verified as

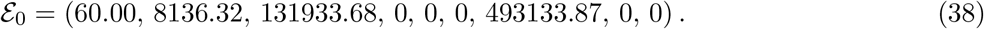

The infinity and Euclidean norms of the model vector field evaluated at *E*_0_ were 7.99 *×* 10^*−*15^ and 1.07 *×* 10^*−*14^, respectively. These near-zero residuals confirm that the computed state satisfies the equilibrium equations to numerical precision.

As an independent consistency check, the infected-state Jacobian was extracted from the complete nine-dimensional nonlinear model by numerical differentiation. For the infected subsystem (*E*_*h*_, *I*_*h*_, *E*_*m*_, *I*_*m*_), the resulting matrix was

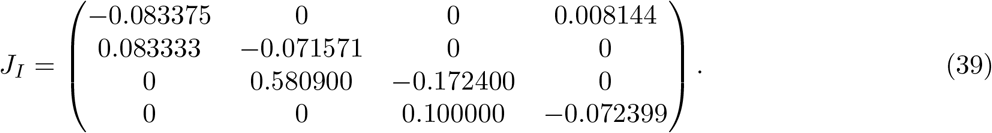

The numerically extracted infected-state Jacobian agreed exactly with the theoretical matrix *F − V* . Both the Frobenius norm of *J*_*I*_ *−* (*F − V*) and the maximum element-wise discrepancy were zero. This exact agreement confirms the consistency of the infected-compartment ordering, force-of-infection terms, and transition structure between the analytical derivation and the implemented nonlinear system.

The directional transmission components were

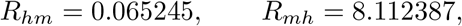

such that

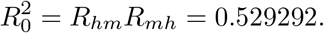

Hence,

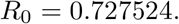

The analytical reproduction number and the spectral radius of the NGM were identical to numerical precision. Since *R*_0_ *<* 1, the DFE is locally asymptotically stable under the reference autonomous conditions. This was independently supported by the infected-state Jacobian, whose spectral abscissa was −1.28148 *×* 10^*−*2^. The full nine-dimensional Jacobian had a spectral abscissa of *−*4.21496 *×* 10^*−*5^. Its considerably smaller magnitude indicates a slowly decaying mode in the wider demographic–awareness system. Thus, although infection-related perturbations may initially decay, convergence of the complete model to the DFE can occur over a substantially longer time scale.

#### 4.5.1 Critical Biting Rate and Stability Threshold

The mosquito biting rate was identified as the most influential transmission parameter in the sensitivity analysis and was therefore selected as the bifurcation parameter for threshold verification. Solving the analytical expression for the basic reproduction number under the condition *R*_0_ = 1 yielded a critical reference biting rate of

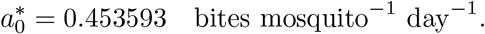

An independent numerical analysis based on the infected-subsystem Jacobian identified the stability transition at 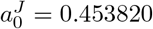, corresponding to *R*_0_ = 1.000501. The relative difference between the analytical and numerical critical biting rates was approximately 0.05%, demonstrating excellent agreement between the next-generation matrix formulation and the Jacobian eigenvalue analysis. This agreement is consistent with the next-generation matrix theory of epidemic models, which predicts that the dominant eigenvalue of the infected subsystem changes sign precisely when the basic reproduction number crosses unity, thereby determining the local stability of the disease-free equilibrium [9, 21].

Figure 6 shows that the basic reproduction number increases monotonically with the reference mosquito biting rate and crosses the epidemic threshold *R*_0_ = 1 at the critical value 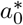. This behaviour agrees with classical malaria transmission theory, in which the mosquito biting rate directly governs the force of infection and is therefore one of the principal determinants of disease persistence [11, 18, 20]. It is also consistent with comparative sensitivity studies showing that mosquito biting rate is among the most influential parameters across a wide range of malaria transmission models [7, 22].

**Fig 6.**
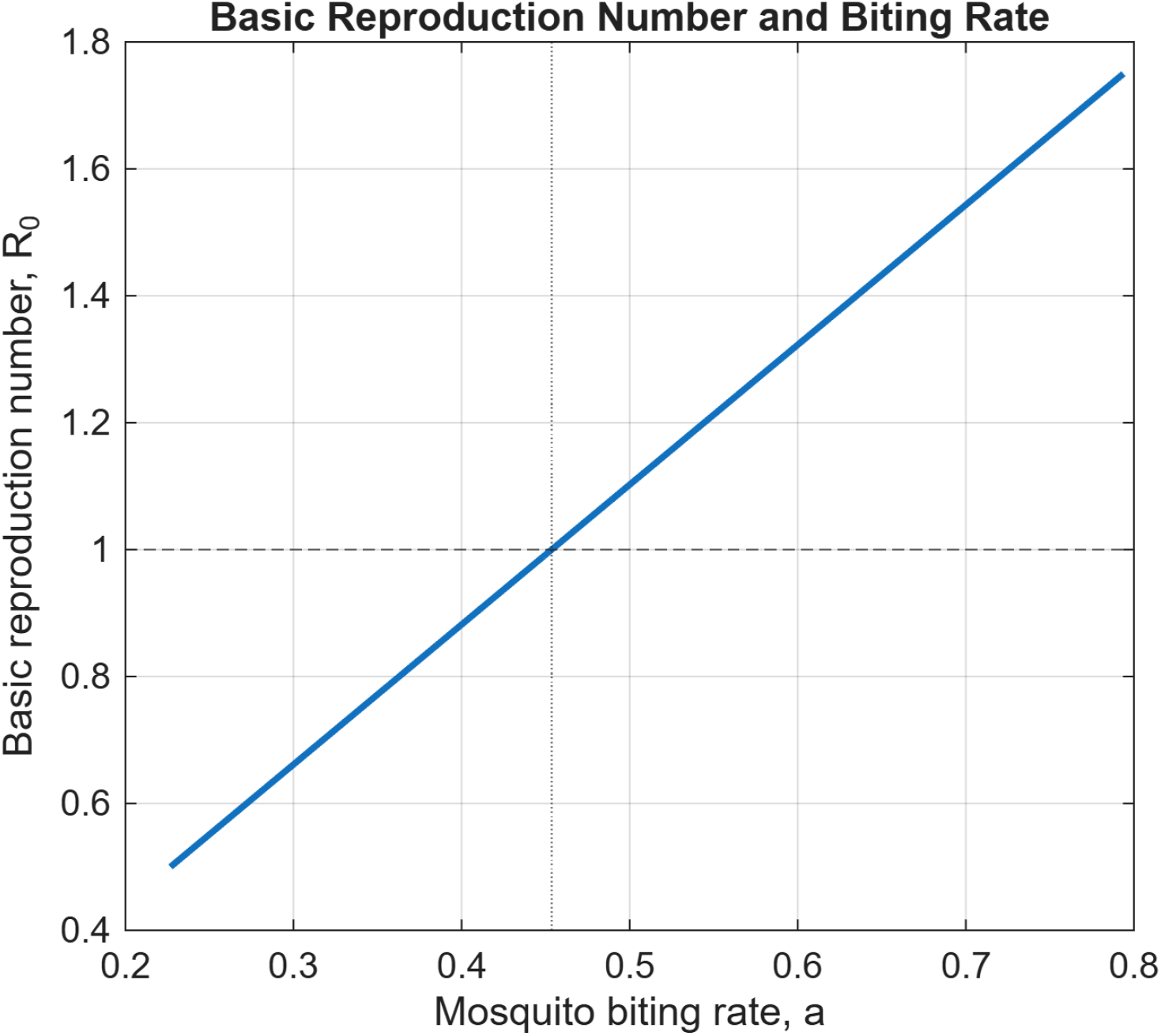
Variation of the basic reproduction number with the reference mosquito biting rate *a*_0_. The horizontal dashed line denotes the epidemic threshold *R*_0_ = 1, while the vertical dashed line identifies the critical biting rate 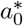.

Figure 7 shows the corresponding spectral abscissa of the infected-state Jacobian. The dominant eigenvalue remains negative for *R*_0_ *<* 1, approaches zero at the threshold, and becomes positive for *R*_0_ *>* 1, confirming the transition from local asymptotic stability to instability of the malaria-free equilibrium. Similar threshold behaviour has been observed in previous malaria modeling studies, where bifurcation analyses with respect to mosquito biting rate and mosquito mortality identified the same stability transition at the epidemic threshold [7, 22].

**Fig 7.**
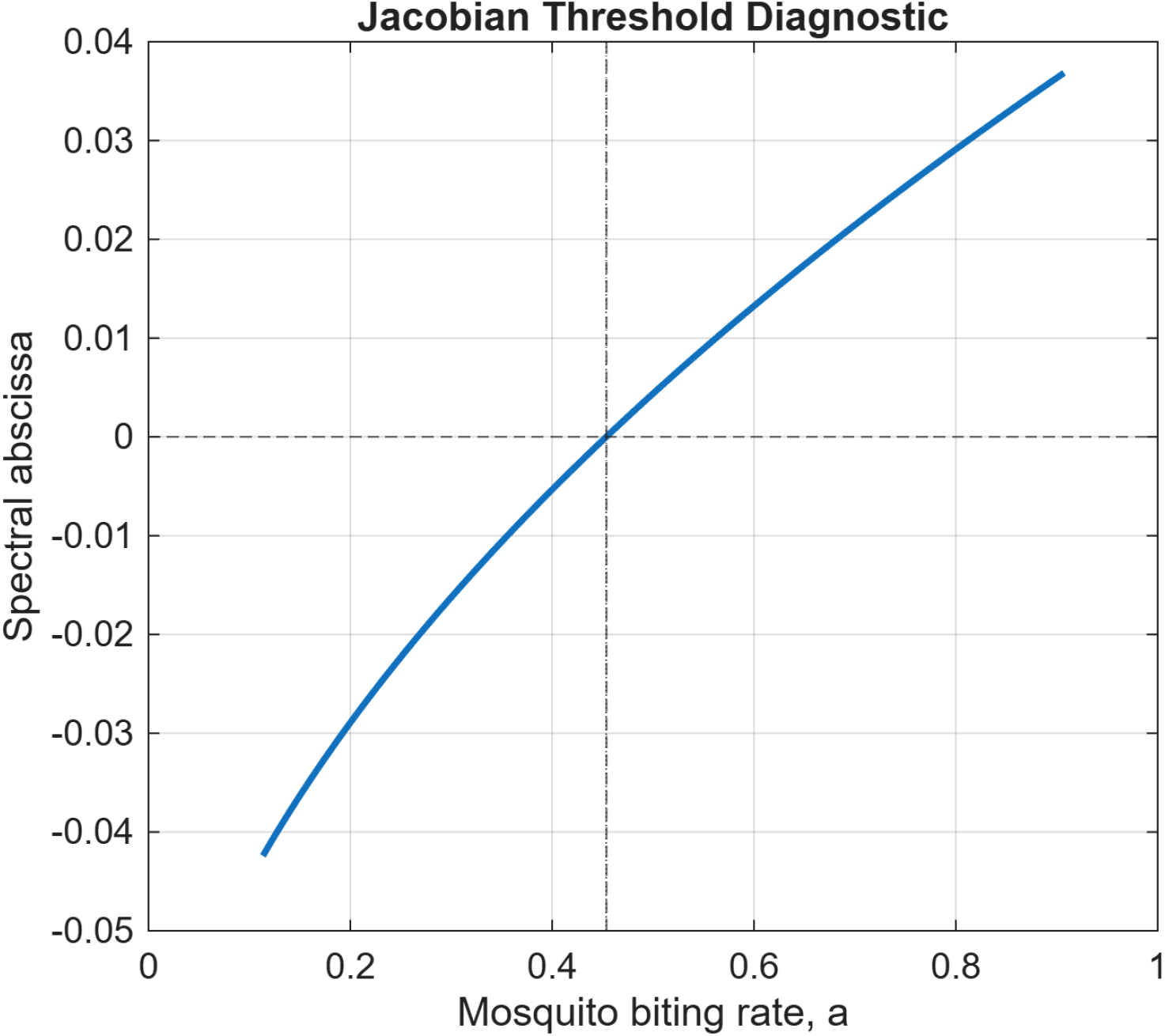
Spectral abscissa, *s*(*J*_*I*_), of the infected-state Jacobian as a function of the reference mosquito biting rate *a*_0_. The zero crossing coincides with the critical threshold *R*_0_ = 1, confirming the local stability result.

Together, Figures 6 and 7 provide complementary numerical verification that the epidemic threshold occurs precisely at *R*_0_ = 1, confirming the theoretical predictions of the next-generation matrix framework. From a public health perspective, the results indicate that interventions capable of reducing mosquito biting rates below the critical threshold—such as long-lasting insecticide-treated nets, indoor residual spraying, and community-based behavioural measures—can shift the dominant eigenvalue into the negative half-plane and thereby prevent sustained malaria transmission [19, 24].

#### 4.5.2 Numerical Verification of the Epidemic Threshold

The epidemic threshold was further verified by varying the mosquito biting rate to generate reproduction numbers in the range 0.5 *≤ R*_0_ *≤* 1.5. Table 7 summarizes the corresponding spectral abscissa and infection outcomes.

**Table 7.**
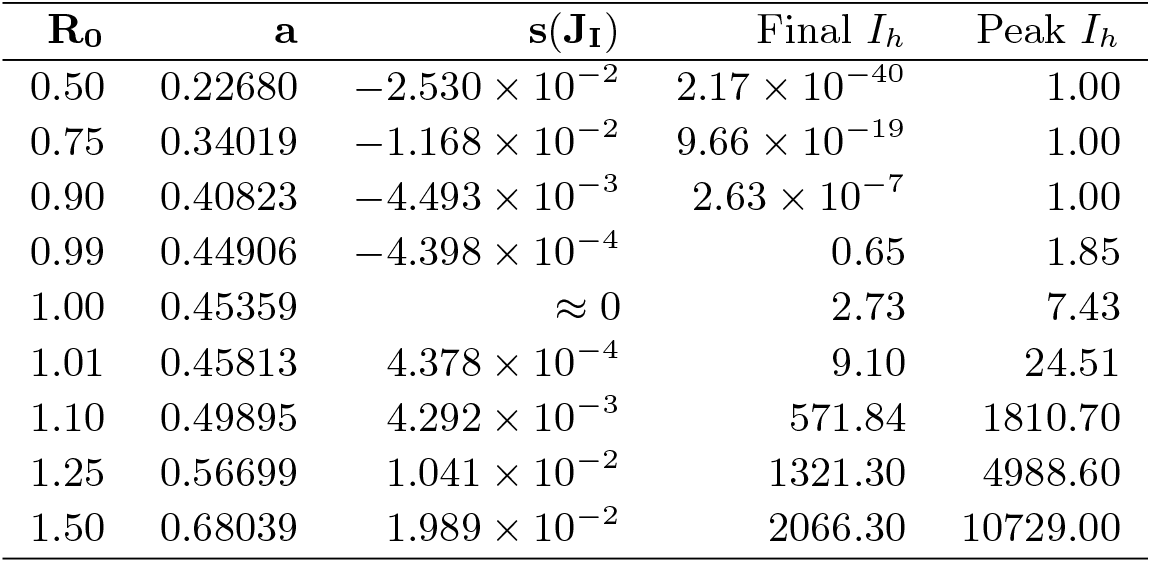
Numerical verification of the epidemic threshold at *R*_0_ = 1.

The results show that the dominant infected-state eigenvalue remained negative for *R*_0_ *<* 1. This indicated that both the local asymptotic stability of the MFE, and the infectious human population, decayed to zero. As *R*_0_ approached unity, convergence became increasingly slow. At *R*_0_ = 0.99, for example, *s*(*J*_*I*_) = *−*4.398 *×* 10^*−*4^ and a small residual infection remained at the end of the finite simulation period. This transient behaviour reflects slow convergence to the disease-free equilibrium rather than endemic persistence.

At the critical threshold *R*_0_ = 1, the dominant eigenvalue was approximately zero, confirming the theoretical bifurcation point. For *R*_0_ *>* 1, the spectral abscissa became positive, and both the equilibrium and peak infectious populations increased rapidly. For example, the peak infectious mhuman population rose from approximately 25 individuals at *R*_0_ = 1.01 to more than 10 700 at *R*_0_ = 1.50, demonstrating the high sensitivity of malaria transmission to small increases in *R*_0_ above unity.

The agreement between the next-generation matrix analysis, Jacobian stability analysis, and long-term numerical simulations allows for sound numerical confirmation that *R*_0_ = 1 is the critical threshold governing malaria persistence in the proposed model.

#### 4.5.3 Equilibrium Continuation and Bifurcation Behaviour

To characterise finite-time persistence from genuine endemic equilibrium, an autonomous frozen-climate equilibrium continuation analysis was performed. A positive endemic equilibrium was first established at *R*_0_ = 1.75 and subsequently tracked backwards toward the threshold. The high-transmission seed was verified with 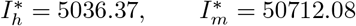,and an equilibrium residual of 3.64 *×* 10^*−*12^. Thus, the initial continuation state represented a genuine positive equilibrium rather than a slowly evolving transient state. The continuation procedure verified all 300 equilibrium points considered over 0.5 *≤ R*_0_ *≤* 1.75, of which 180 were positive endemic states. The first verified endemic equilibrium occurred at (*R*_0_ = 1.001672, *a* = 0.454352). This threshold is immediately above unity and closely agrees with both the analytical critical biting rate *a*_*c*_ = 0.453593 and the Jacobian estimate 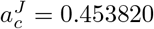.

Near the threshold, the endemic branch approached the DFE continuously. Specifically, 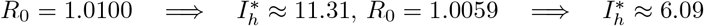, and 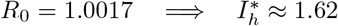.

At the next continuation point below unity, *R*_0_ = 0.9975, no positive endemic equilibrium was verified.

More generally, no endemic branch was detected for *R*_0_ *<* 1.

Figure 8 shows the equilibrium infectious human population as a function of *R*_0_. The positive endemic branch emerges continuously from the neighbourhood of the DFE immediately above unity and increases with transmission intensity. The absence of a verified positive branch below unity and the continuous emergence of the endemic equilibrium above the critical point are consistent with a forward transcritical bifurcation.

**Fig 8.**
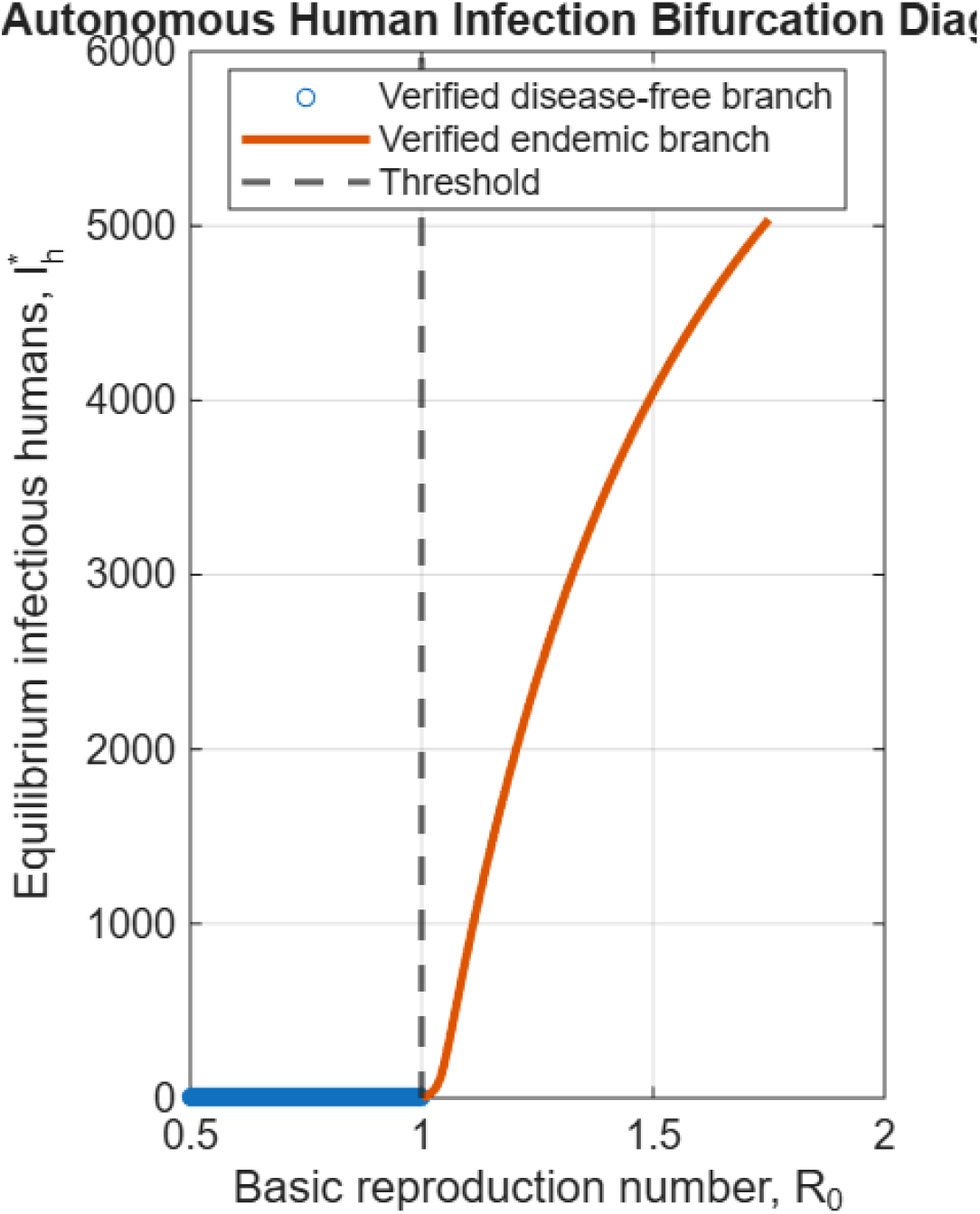
Bifurcation diagram showing the equilibrium infectious human population 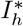 as a function of *R*_0_. The positive endemic branch emerges immediately above *R*_0_ = 1, consistent with a forward transcritical bifurcation.

The automatic numerical classification therefore identified a *forward bifurcation*. This finding has an important control interpretation. In a system exhibiting backward bifurcation, reducing *R*_0_ below unity may be insufficient for elimination because a stable endemic state can coexist with the DFE. No such behaviour was detected under the parameter regime considered here. Instead, the equilibrium continuation results support the classical threshold criterion whereby sustained reduction of the effective reproduction number below unity ultimately drives the autonomous system toward the disease-free state.

#### 4.5.4 Critical Slowing Down and Epidemiological Implications

A notable feature of the threshold analysis was the pronounced slowing of epidemic decay as *R*_0_ approached unity from below. The infected-state spectral abscissa increased from −2.530 *×* 10^*−*2^ at *R*_0_ = 0.50 to −4.398 *×* 10^*−*4^ at *R*_0_ = 0.99. Although the DFE remained locally asymptotically stable for *R*_0_ *<* 1, the characteristic rate of return to the disease-free state therefore decreased substantially near the threshold.

This behaviour is consistent with critical slowing down. A malaria system with *R*_0_ only slightly below unity may retain detectable infection for a prolonged period and accumulate considerable burden before eventual elimination. The finite-time persistence observed at *R*_0_ = 0.99 is therefore compatible with DFE stability and is fundamentally different from the positive equilibrium branch verified for *R*_0_ *>* 1.

The result also clarifies the relationship between the autonomous threshold analysis and the seasonally forced simulations. The estimate *R*_0_ = 0.727524 characterizes invasion under frozen reference climatic conditions and does not imply that transmission potential remains subcritical throughout a season.

Climate-driven variation in biting, mosquito-to-human transmission probability, mosquito recruitment, mortality, and extrinsic incubation can temporarily increase vectorial capacity. Such periods may amplify infection even when the reference autonomous system is subcritical.

From a control perspective, maintaining *R*_0_ only marginally below unity may therefore provide insufficient short-term protection against substantial malaria burden. As the threshold is approached, epidemic decay slows and small climate- or behaviour-induced changes in transmission can move the system into the supercritical regime. Intervention strategies should thus seek to maintain transmission sufficiently below the critical boundary rather than treating *R*_0_ = 1 as a narrow operational target.

Overall, the exact Jacobian–NGM agreement, the close correspondence between analytical and numerical critical biting rates, the sign change of the dominant infected-state eigenvalue, and the equilibrium-verified forward endemic branch provide a coherent set of independent validations of the model threshold. These results establish *R*_0_ = 1 as the central invasion boundary of the autonomous system and provide the dynamical basis for the subsequent evaluation of time-dependent optimal intervention strategies.

### 4.6 Optimal Control Simulation Results

The numerical optimal-control problem was solved over a 365-day intervention horizon using the forward-backward sweep method. The four time-dependent controls represented long-lasting insecticidal net (LLIN) use, awareness intervention, indoor residual spraying (IRS), and prompt treatment, respectively. The state weights assigned to (*E*_*h*_, *I*_*h*_, *E*_*m*_, *I*_*m*_) were 1.0, 5.0, 0.05, and 0.10, while the quadratic control-cost weights for LLINs, awareness, IRS, and treatment wer 500, 100, 1000, and 100, respectively. Before solving the optimal-control problem, the consistency of the implemented Hamiltonian derivatives was examined numerically. All four controls produced non-zero changes in the state vector field, confirming that each intervention was active in the implemented model. Furthermore, the direct Hamiltonian control gradients agreed with independent finite-difference approximations. The maximum absolute and relative discrepancies were 5.78 *×* 10^*−*8^ and 1.08 *×* 10^*−*9^, respectively. These small errors provide strong computational evidence that the control derivatives used in the optimization were consistent with the implemented state equations.

The combined four-control forward-backward sweep converged after 25 iterations with a final convergence error of 6.33 *×* 10^*−*7^. The terminal adjoint infinity norm was zero, all optimal controls remained in the admissible interval [0, 1], and the maximum projected-gradient residual was 1.23 *×* 10^*−*4^. The numerical solution therefore satisfied the principal convergence, transversality, admissibility, and first-order optimality diagnostics.

#### 4.6.1 Effect of Optimal Control on Human Infection

Under the uncontrolled scenario, the cumulative infectious-human burden was 40 038.97 person-days and the peak infectious-human population was 193.99. Application of the optimal four-control strategy reduced the cumulative human infection burden by 92.49% and the peak infectious-human population by 47.54%.

Figure 9 compares the infectious-human trajectories under the uncontrolled and optimally controlled scenarios. The uncontrolled trajectory exhibits a pronounced epidemic increase followed by a gradual decline. In contrast, the optimally controlled trajectory is substantially suppressed throughout the intervention horizon. The difference between the two trajectories is particularly pronounced during the period of highest transmission pressure, demonstrating that optimal intervention reduces both the magnitude and duration of human infection.

**Fig 9.**
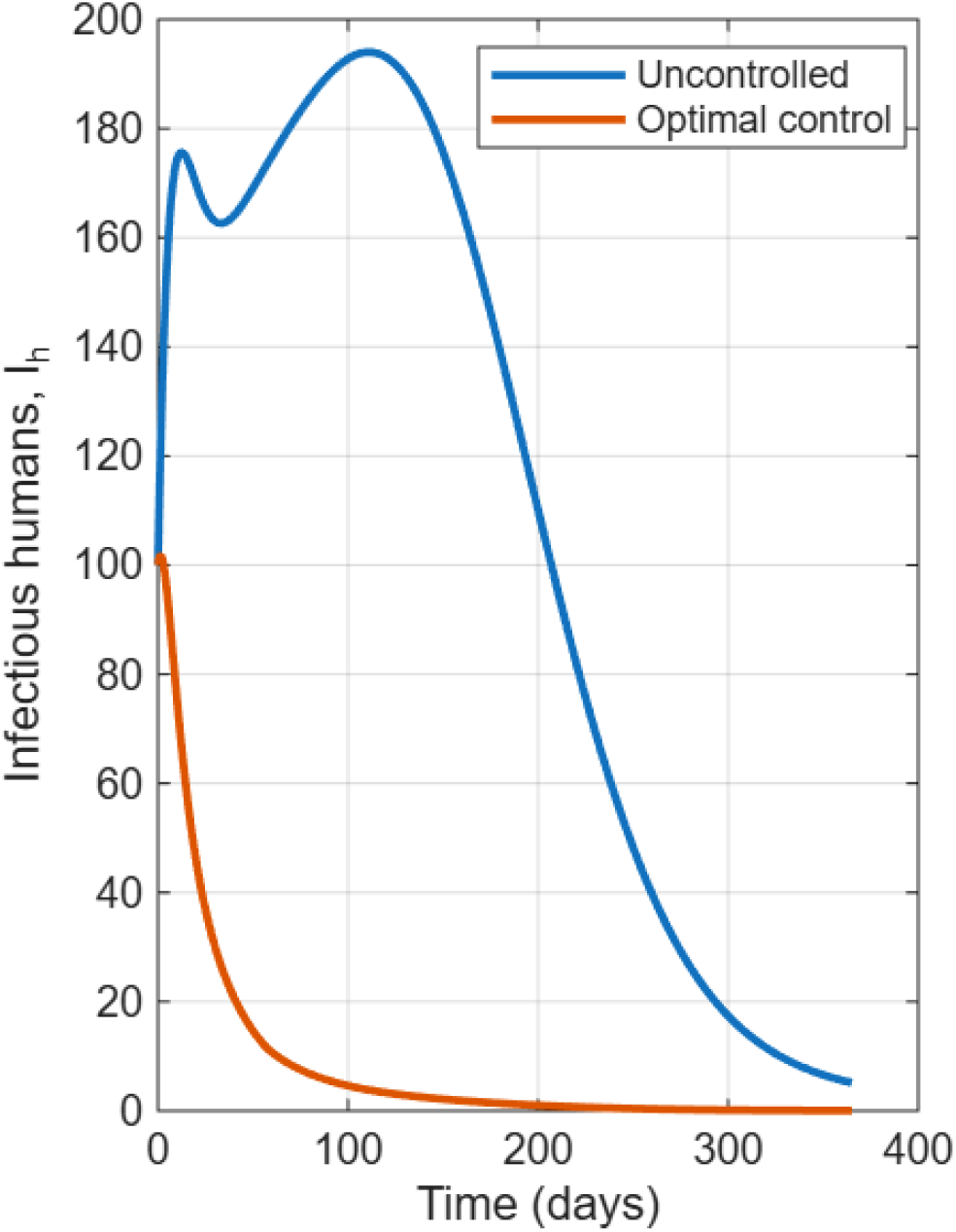
Infectious-human trajectories under uncontrolled and optimal four-intervention strategies. Optimal control substantially suppresses human infection and reduces cumulative infectious-human burden by 92.49%.

The reduction in cumulative burden was substantially larger than the reduction in peak prevalence. This distinction indicates that the principal benefit of optimal intervention was not limited to lowering the epidemic maximum. Rather, the controls substantially shortened the duration for which large numbers of infectious individuals remained in the population. Consequently, the integrated disease burden declined by more than 92% despite a peak reduction of approximately 48%.

This result is epidemiologically important because cumulative infectious burden reflects both infection magnitude and persistence. A strategy that moderately reduces the epidemic peak but rapidly removes infectious individuals can generate a substantially larger reduction in total transmission opportunity than would be inferred from peak prevalence alone.

#### 4.6.2 Effect of Optimal Control on Mosquito Infection

The uncontrolled cumulative infectious-mosquito burden was 230 819.72 mosquito-days, with a peak infectious-mosquito population of 1272.58. Under the combined optimal-control strategy, cumulative mosquito burden declined by 93.87%, while peak infectious-mosquito abundance was reduced by 80.00%.

The corresponding mosquito infection trajectories are presented in Figure 10. Optimal intervention produced a marked reduction in the infectious-vector population throughout the simulation period. The substantial suppression of *I*_*m*_ limits the reservoir of infectious mosquitoes capable of transmitting malaria to susceptible humans and therefore weakens the mosquito-to-human component of the transmission cycle.

**Fig 10.**
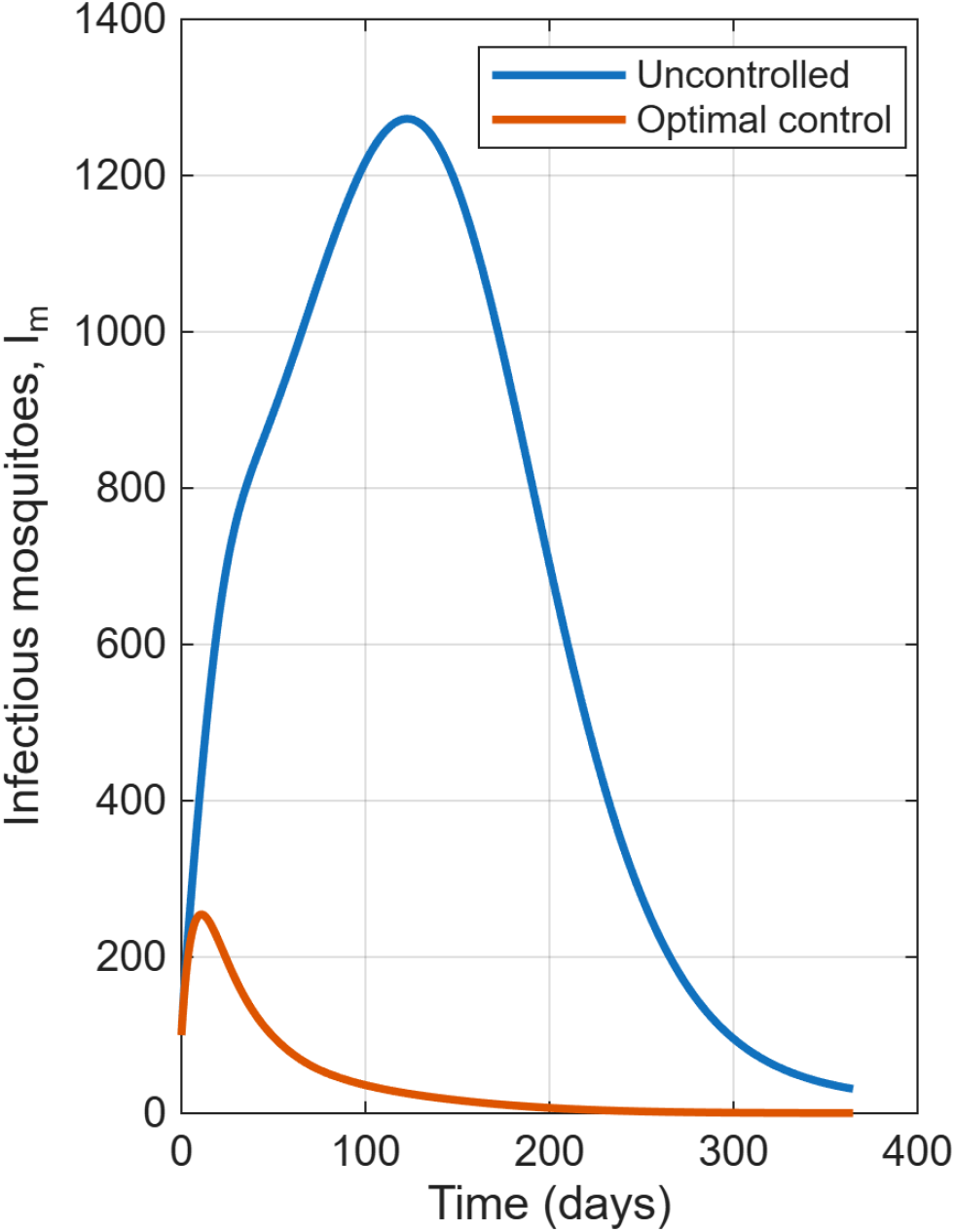
Infectious-mosquito trajectories under uncontrolled and optimal control scenarios. The combined optimal strategy reduces cumulative infectious-mosquito burden by 93.87% and peak infectious-mosquito abundance by approximately 80.00%.

The simultaneous reduction in human and mosquito infection demonstrates the advantage of targeting both sides of the host–vector transmission cycle. Prompt treatment reduces the duration of human infectiousness, thereby limiting the source of infection available to susceptible mosquitoes. Vector-directed interventions, particularly IRS and LLINs, reduce vector survival or effective contact with humans. The combined strategy therefore generates complementary epidemiological effects that interrupt successive stages of malaria transmission.

#### 4.6.3 Temporal Profiles of the Optimal Controls

The optimal time-dependent intervention profiles are shown in Figure 11. The four controls exhibited substantially different temporal intensities, reflecting differences in their marginal epidemiological benefits and quadratic implementation costs.

**Fig 11.**
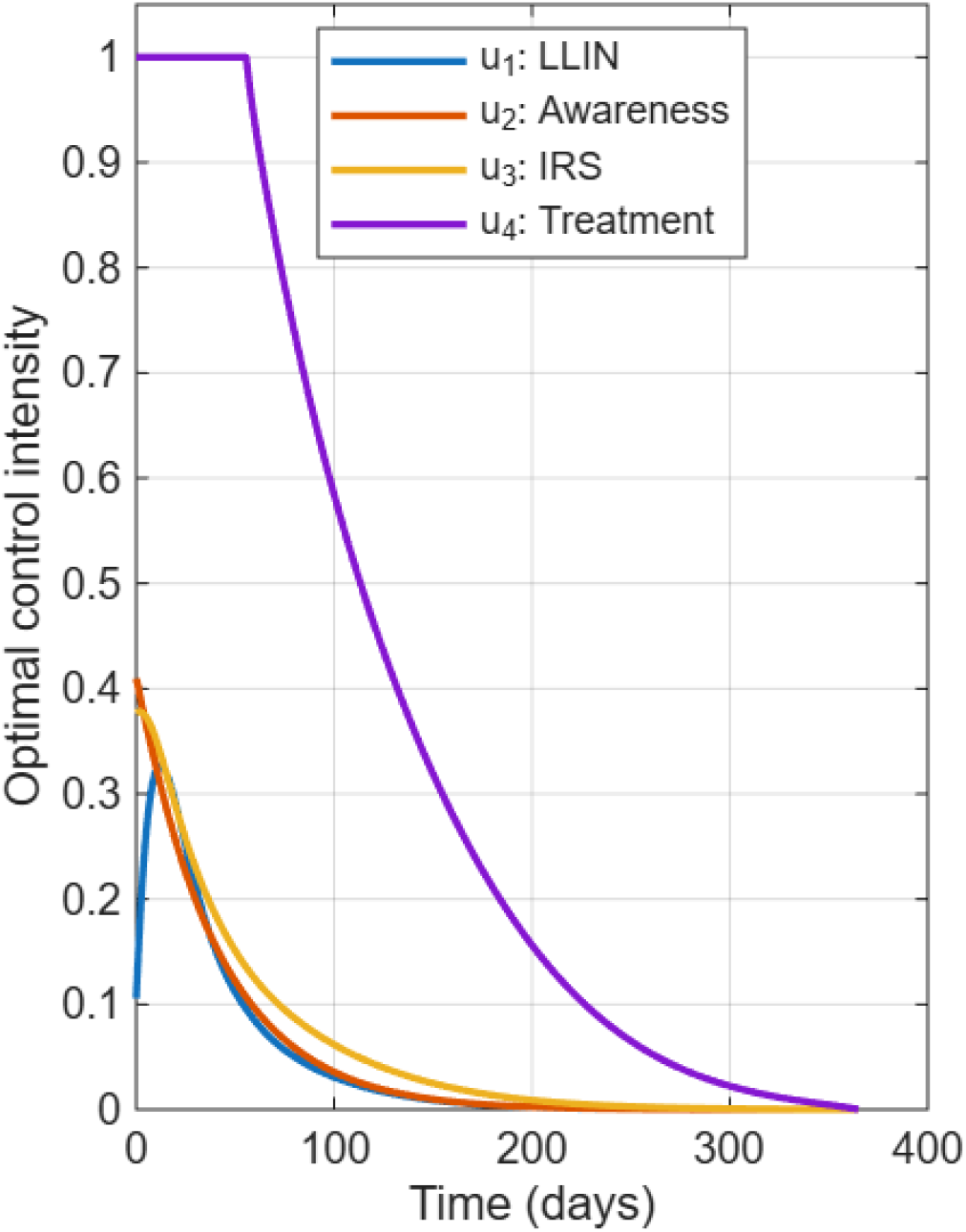
Optimal time-dependent control profiles for LLIN use (*u*_1_), awareness intervention (*u*_2_), indoor residual spraying (*u*_3_), and prompt treatment (*u*_4_).

Prompt treatment was the most intensively deployed intervention. Its mean optimal intensity was 0.3594, and the control reached its upper admissible bound of unity. Treatment remained at or above 90% intensity for approximately 63.25 days. This pattern indicates that rapid removal of infectious humans was strongly favoured during the period of greatest epidemiological benefit.

In comparison, LLIN use had a mean optimal intensity of 0.0425 and a maximum of 0.3266. Awareness had a mean intensity of 0.0461 and a maximum of 0.4097, whereas IRS had a mean intensity of 0.0584 and a maximum of 0.3780. None of these three interventions remained above 90% intensity.

The optimal profiles therefore do not support continuous maximum deployment of every intervention.

Instead, the optimization favours intensive treatment combined with lower, strategically timed levels of LLIN, awareness, and IRS control. This allocation reflects the objective functional used in the analysis, which balances reductions in human and mosquito infection against the quadratic costs of intervention implementation.

The high treatment intensity is also consistent with the relatively large weight assigned to infectious humans in the objective functional. Since *I*_*h*_ received a weight of 5.0, rapid removal of infectious individuals provides a substantial marginal reduction in the objective. Treatment therefore becomes a central component of the optimal intervention programme.

#### 4.6.4 Comparison of Optimal Intervention Strategies

To examine the relative contribution of individual and combined interventions, eleven optimal-control strategies were evaluated. These comprised four single-control strategies, six pairwise intervention combinations, and the full four-control programme. The resulting epidemiological and objective-functional outcomes are summarized in Table 8.

**Table 8.** Comparison of optimal malaria intervention strategies.

| Strategy | Objective | Human burden | Human reduction (%) | Mosquito reduction (%) | Peak $I_h$ reduction (%) |
| --- | --- | --- | --- | --- | --- |
| LLIN | 142890 | 17442.0 | 56.44 | 55.45 | 12.98 |
| Awareness | 215290 | 31942.0 | 20.22 | 18.72 | 9.48 |
| IRS | 109370 | 13171.0 | 67.10 | 88.45 | 10.11 |
| Treatment | 36559 | 3737.0 | 90.67 | 88.66 | 47.53 |
| LLIN + Awareness | 138090 | 17223.0 | 56.98 | 55.64 | 12.98 |
| LLIN + IRS | 94075 | 10521.0 | 73.72 | 88.67 | 12.45 |
| LLIN + Treatment | 34967 | 3408.0 | 91.49 | 89.51 | 47.54 |
| Awareness + IRS | 103210 | 12550.0 | 68.66 | 88.28 | 10.11 |
| Awareness + Treatment | 35998 | 3651.4 | 90.88 | 88.88 | 47.53 |
| IRS + Treatment | 32237 | 3166.1 | 92.09 | 93.82 | 47.53 |
| All controls | 31235 | 3008.3 | 92.49 | 93.87 | 47.54 |

Among the single-intervention strategies, treatment produced the greatest reduction in human disease burden. Treatment alone reduced cumulative human infection by 90.67% and peak infectious-human prevalence by 47.53%. Its objective value of 36 559 was considerably lower than the values obtained for LLINs, awareness, or IRS alone. The result identifies prompt treatment as the dominant single intervention for controlling human infection under the assumed epidemiological and economic weights.

IRS was the strongest single vector-directed intervention. It reduced cumulative infectious-mosquito burden by 88.45% and peak mosquito infection by 84.51%. However, the corresponding reduction in peak human infection was only 10.11%. This result illustrates that strong vector suppression does not necessarily translate immediately into an equivalent reduction in the human epidemic peak, particularly when infectious humans already present in the population continue to contribute to the transmission process.

LLINs produced intermediate reductions of 56.44% and 55.45% in cumulative human and mosquito burdens, respectively. Awareness alone was the least effective strategy under the current parameterization, reducing human burden by 20.22% and mosquito burden by 18.72%. The relatively modest effect of awareness is consistent with its indirect mechanism of action in the model, where behavioural protection operates through movement into the aware susceptible class rather than direct removal of infectious humans or infectious mosquitoes.

Figure 12 compares the cumulative human burden reductions obtained under the eleven strategies. Strategies containing treatment consistently produced the largest reductions. The addition of IRS to treatment generated a further improvement, producing a 92.09% reduction in cumulative human burden.

**Fig 12.**
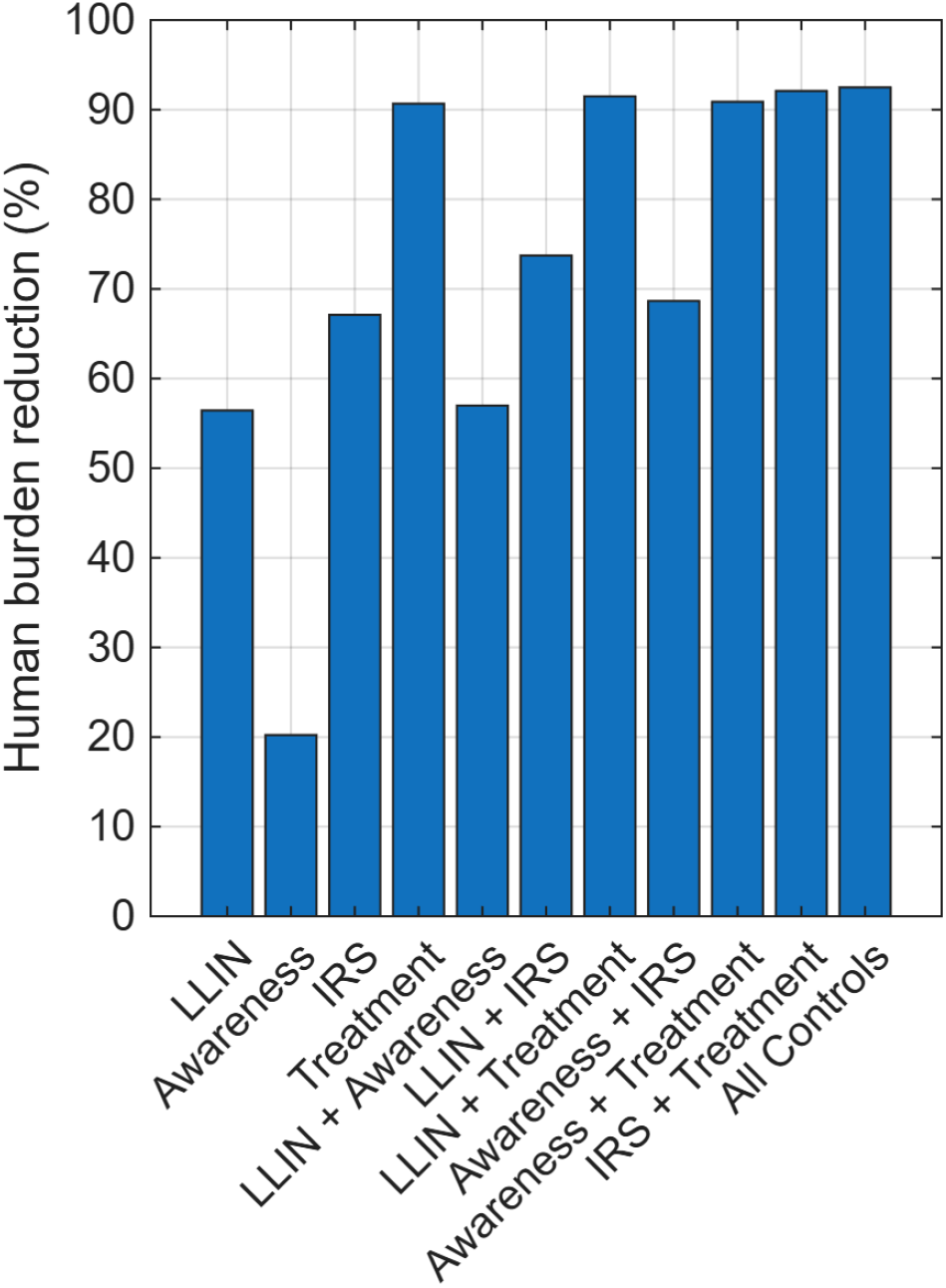
Percentage reduction in cumulative infectious-human burden under alternative optimal-control strategies. Treatment-containing strategies produce the largest reductions, with the full four-control strategy achieving the maximum reduction of 92.49%.

A particularly important result is the near-optimal performance of the **IRS + Treatment** strategy. This combination reduced cumulative human burden by 92.09% and cumulative mosquito burden by 93.82%, compared with 92.49% and 93.87%, respectively, under the full four-control programme. Thus, adding LLIN and awareness controls to the IRS–treatment strategy increased human burden reduction by only approximately 0.39 percentage points and mosquito burden reduction by approximately 0.05 percentage points.

The objective value for IRS + Treatment was 32 237, compared with 31 235 for the full strategy. The relative difference in the objective functional was therefore modest. These findings suggest that the combination of prompt case management and vector suppression captures most of the epidemiological benefit obtained from simultaneous implementation of all four interventions.

This near-optimality has potential resource-allocation implications. Where financial, logistical, or operational constraints limit the simultaneous deployment of multiple malaria interventions, prioritizing treatment and IRS may provide a highly efficient alternative to the full intervention package. However, this interpretation is conditional on the control-cost weights and parameterization adopted in the present analysis and should not be viewed as evidence that LLINs or awareness interventions are epidemiologically unimportant in all settings.

#### 4.6.5 Reduction in Peak Human Infection

Figure 13 presents the percentage reduction in peak infectious-human prevalence under the alternative intervention strategies. A distinct separation was observed between treatment-containing and non-treatment strategies.

**Fig 13.**
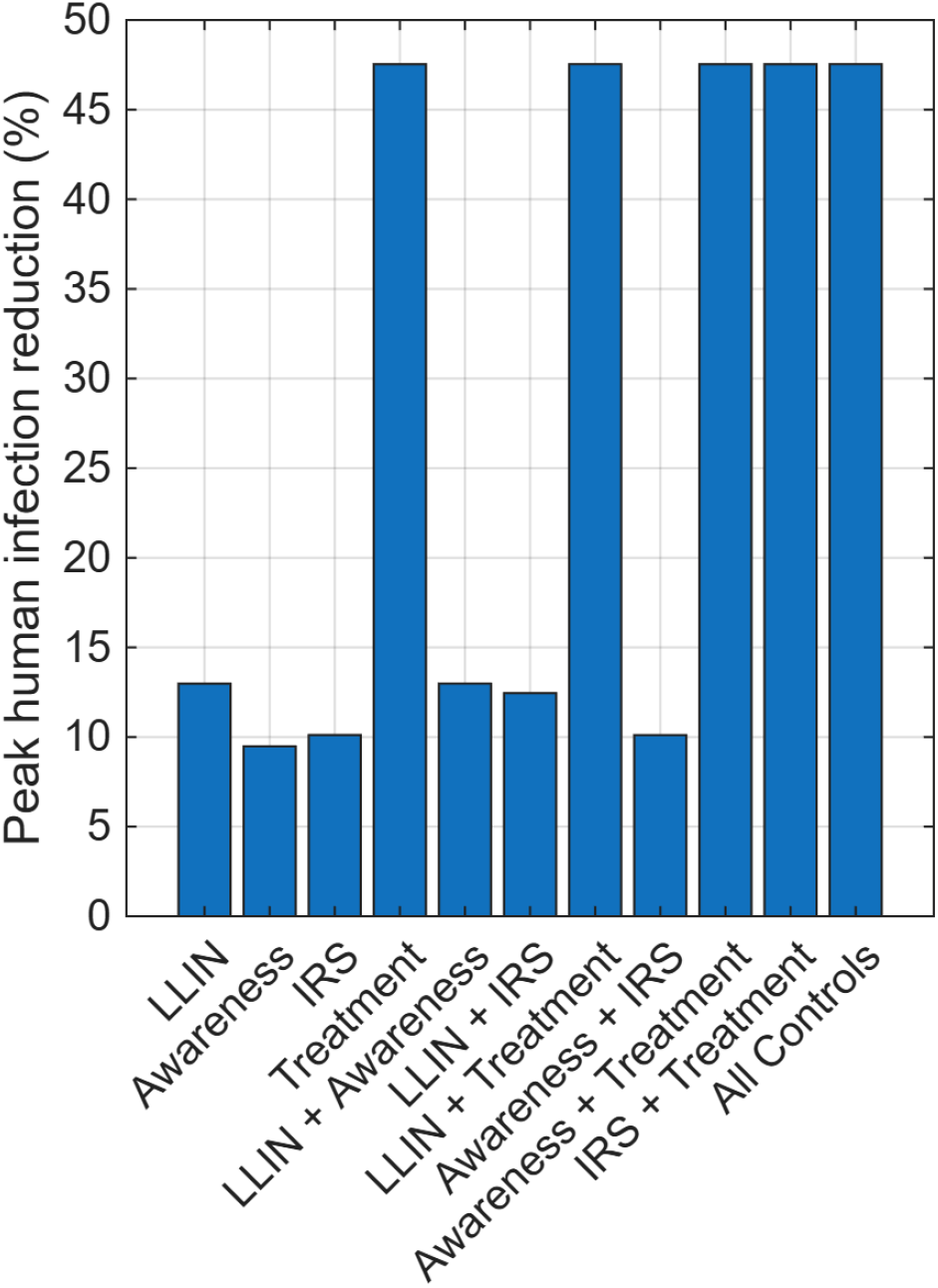
Percentage reduction in peak infectious-human prevalence under alternative optimal-control strategies. Strategies containing prompt treatment achieve substantially greater peak reduction than interventions without treatment.

Treatment alone reduced peak human infection by 47.53%. The addition of LLINs, awareness, IRS, or all controls produced only small additional changes in this endpoint, with the full strategy achieving a peak reduction of 47.54%. By contrast, the peak reductions associated with LLIN, awareness, and IRS alone were 12.98%, 9.48%, and 10.11%, respectively.

The results indicate that the human epidemic peak is principally controlled through the rapid removal of infectious individuals in the present model. Vector-control interventions generate substantial reductions in cumulative transmission and mosquito infection but have a more gradual influence on the maximum number of infectious humans. Consequently, treatment dominates the short-term reduction of peak human prevalence, whereas IRS contributes strongly to longer-term transmission suppression.

Interestingly, IRS + Treatment achieved a peak mosquito reduction of 80.60%, compared with 80.00% under the full four-control strategy. This does not contradict the optimality of the full strategy because the objective functional minimizes a weighted integral of infection states and control costs rather than any single epidemiological endpoint. A strategy may therefore perform slightly better for a specific outcome while having a larger overall objective value.

#### 4.6.6 Convergence of the Optimal-Control Algorithm

The convergence behaviour of the combined four-control forward–backward sweep is shown in Figure 14. The convergence error declined rapidly during the initial iterations and continued to decrease approximately monotonically until the prescribed tolerance was satisfied.

**Fig 14.**
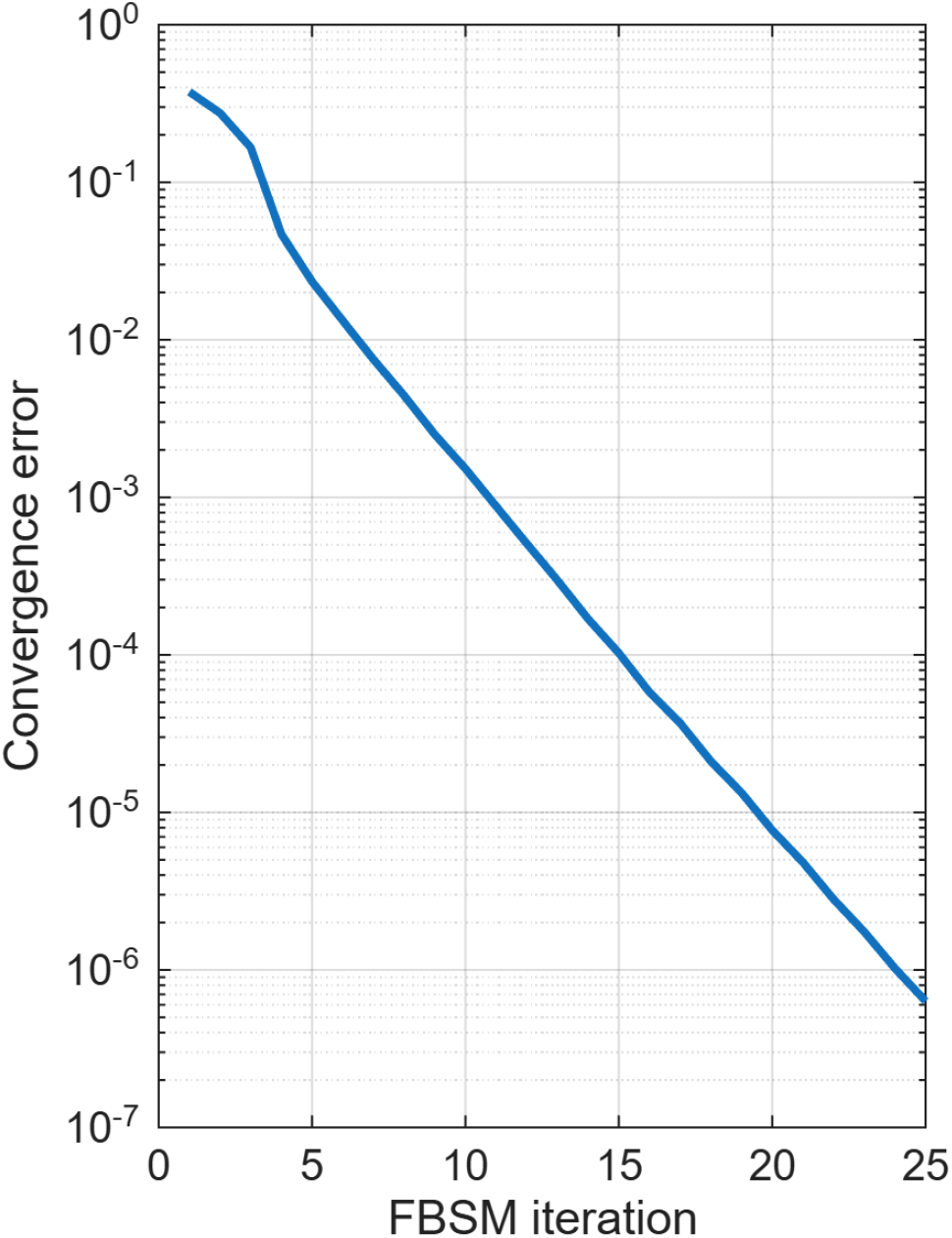
Convergence history of the forward–backward sweep algorithm for the combined four-control strategy. The algorithm converged after 25 iterations with a final error of 6.33 *×* 10^*−*7^.

The combined strategy converged after 25 iterations. All eleven strategy optimizations also converged successfully, requiring between 18 and 32 iterations. The IRS-only strategy converged most rapidly in 18 iterations, whereas LLIN + Awareness required 32 iterations. No strategy reached the maximum iteration limit. The convergence results, together with the zero terminal adjoint residual, control-bound verification, Hamiltonian-gradient consistency test, and small projected-gradient residual, support the numerical reliability of the computed optimal-control solutions.

## 5 Overall Epidemiological and Control Implications

The optimal-control simulations revealed that the combination of various malaria interventions leads to a significant reduction in disease transmission, infection levels, and overall cost of control measures. Compared to the scenario without any control interventions, the simultaneous use of LLINS, awareness campaigns, IRS, and prompt treatment reduced the objective functional by 88.17%, while the total number of human infections and infectious mosquitoes was lower by 92.49% and 93.87%, respectively. This result highlights the tremendous impact of synergy in malaria control programs and demonstrates that the combination of various control strategies can have a significantly better effect than the use of individual interventions.

The numerical results also revealed that the contribution of each intervention to the overall reduction of disease transmission is different. While prompt treatment reduces the length of the infectious period, IRS decreases the number of infectious mosquitoes due to the lower survival rates. LLINS reduce the biting rate and, consequently, the force of infection, whereas awareness campaigns increase overall compliance with prevention measures and promote treatment-seeking behavior. The combined effect of these measures leads to an even greater reduction in disease prevalence because each intervention targets different links in the transmission cycle.

An interesting observation from the optimal control simulations is that the combined use of IRS and treatment was almost as effective as the combination of all four interventions. Although the simultaneous use of all malaria interventions resulted in the greatest decreases in prevalence and the lowest costs, the additional benefit from the use of LLINS, awareness campaigns, and prompt treatment over IRS and treatment was relatively modest. This result has important implications for the allocation of resources towards malaria control in low-income endemic regions since even a small reduction in prevalence is critical for areas with limited healthcare resources. Thus, IRS combined with prompt diagnosis and treatment may be an adequate strategy for many malaria-endemic regions with limited healthcare infrastructure.

The numerical experiments also illustrate the theoretical results of this study. The numerical simulations of the model with control interventions demonstrated that the reduction in the number of infectious mosquitoes and the decrease in the force of infection lead to a rapid decline in prevalence. The stability analysis revealed that the malaria-free equilibrium is locally stable when *R*_0_ *<* 1 and becomes unstable when *R*_0_ *>* 1, which implies that a malaria outbreak can only occur when *R*_0_ *>* 1. In addition, the optimal control strategies reduced _0_ below the critical threshold, causing a forward bifurcation that decreased the prevalence of the disease.

From an epidemiological perspective, the results of this study highlight the importance of using a combination of vector- and host-targeted measures to efficiently reduce the prevalence of malaria. Vector control, in the form of IRS and LLINS, reduces the number of infectious mosquitoes and the biting rate, thus decreasing the force of infection. Prompt treatment lowers the proportion of infectious humans and shortens the duration of the infectious period. The inclusion of awareness into the model demonstrates that effective behavioral change can significantly increase the efficacy of existing control measures. The role of awareness campaigns in reducing the spread of the disease illustrates the importance of community engagement in any large-scale public health campaign. This is especially important for regions with seasonal climatic conditions since community participation can significantly affect the epidemiological situation with malaria in these areas.

The results of this study also have important implications for policymakers. If sufficient funding is available, the simultaneous use of all four interventions is the preferred option, as it leads to the greatest decreases in prevalence. However, in regions with limited healthcare resources, the allocation of funds towards IRS and prompt treatment has the best cost-benefit ratio, as illustrated by the optimal control results. Thus, the results of this study can be used in policymaking to determine the most plausible allocation of resources towards malaria control in a region based on its healthcare situation.

In conclusion, this study demonstrated that the combination of various mathematical and computational methods can be useful in determining the most effective strategies for malaria control in different regions. The proposed model highlights the importance of vector control and behavioral change in reducing the prevalence of the disease and can be useful in policymaking in malaria-endemic regions.

## Data Availability

Data used in the study was majorly simulated data. The Matlab code used will be made available once required.

